# Heterogeneity of Treatment Effect of Aspirin and Clinically Significant Bleeding in Older Adults

**DOI:** 10.64898/2026.06.10.26355385

**Authors:** Giorgos Tzimas, Roselyne B. Tchoua, Joseph C. Vanghelof, Rory Wolfe, Geoffrey Cloud, Suzanne E. Mahady, Lianlian Du, Michael E. Ernst, Erica M. Wood, Daniela S. Raicu, Shara Ket, Raj C. Shah

**Author notes:** (Corresponding Author) Raj C. Shah. **Author Contact Information:** Giorgos Tzimas, Roselyne B. Tchoua, Joseph C. Vanghelof, Daniela S. Raicu, Michael E. Ernst, Erica M. Wood, Geoffrey Cloud, Suzanne E. Mahady, Rory Wolfe, Shara Ket, Lianlian Du.

## Abstract

**Aim:** The global population of older adults is growing, and older age is linked to higher bleeding risk. Although guidelines discourage aspirin for primary prevention in healthy older adults due to bleeding harms outweighing benefits, many continue taking it without a clear indication. It remains unclear whether all older adults face uniform aspirin-related bleeding risk or if certain subgroups are more vulnerable.

**Methods:** We analyzed data from 19,114 ASPREE trial participants to develop machine learning models using 116 baseline variables. Random forest (RF) and random survival forest (RSF) models predicted 5-year bleeding risk, and participants were stratified into low, intermediate, and high-risk groups based on the 20th and 80th percentiles of predicted risk. We assessed heterogeneity of treatment effect (HTE) by testing treatment-by-risk group interactions on the relative scale using Fine-Gray models, and on the absolute scale using observed 5-year cumulative incidence rates.

**Results:** Over a median follow-up of 4.7 years, 626 major bleeding events occurred. The RF model had moderate discrimination (AUC = 0.65, 95% CI: 0.63-0.67) and good calibration (Brier = 0.032, 95% CI: 0.029-0.034). Statistically significant HTE was observed on the relative scale, with the greatest relative increase in bleeding risk seen in the low-risk group (subdistribution hazard ratio = 2.26, 95% CI: 1.27–4.01). On the absolute scale, low-risk participants experienced higher bleeding with aspirin (absolute risk difference (ARD) = 1.17%, 95% CI: 0.37–1.95), but heterogeneity in ARDs was not statistically significant (Cochran’s Q p > 0.45). Similar findings were observed when using the RSF model.

**Conclusion:** Participants at lowest baseline bleeding risk experienced the greatest relative increase in bleeding risk with aspirin therapy. We found statistically significant heterogeneity in treatment effects on the relative but not absolute scale. These findings support an individualized, risk-based approach to aspirin therapy decision-making in older adults.

## Introduction

The global population of older adults is experiencing rapid growth, driven by significant improvements in living standards, advances in medical care, and public health measures(1). The probability of experiencing a major bleeding event increases with age and is recognized as a significant cause of both mortality and morbidity(2). This risk is further increased by the use of certain drug therapies such as aspirin, a commonly used antiplatelet agent known to increase bleeding risk through inhibition of platelet aggregation(3,4). In 2016, the U.S. Preventive Services Task Force (USPSTF) recommended against the use of aspirin for primary prevention in adults aged 70 years and older due to a poor balance of benefits and harms(5). In 2022, the USPSTF reaffirmed their concerns about bleeding risks related to aspirin use, but their recommendations did not address whether specific subgroups of older adults may face disproportionately higher bleeding risks from aspirin therapy(6). Despite lack of supportive evidence, many older adults continue to take aspirin, often without a clear medical indication(7). Individual characteristics such as comorbidities, lifestyle factors, and demographic characteristics can play a critical role in influencing treatment responses and general susceptibility to bleeding, but aspirin’s benefit/risk in older adults have not been decisively evaluated with these factors considered.

Randomized controlled trials (RCTs) typically assess heterogeneity in treatment effects (HTE) through conventional subgroup analysis throughout the study population(8), where treatment effects are compared between different predefined groups of patients based on a single covariate (such as male vs female)(9). Although this approach is straightforward, easy to interpret, and capable of effectively highlighting these treatment differences, there are several limitations associated with it(10). For example, dividing the patient cohort into multiple levels of a covariate, such as the use of drugs (e.g., the use of anticoagulants, antiplatelets, or other cardiovascular drugs), can significantly reduce the sample size within each subgroup. This could decrease the statistical power needed to detect meaningful differences in treatment effects, especially if those effects are subtle. Furthermore, the risk of false positive findings increases due to the need for multiple comparisons, as each subgroup adds an additional layer of statistical testing, thus increasing the probability of identifying a treatment effect by chance alone(11).

An alternative approach to identifying variability in treatment effect is predictive HTE analysis(12). A common method is risk modeling, in which a multivariate predictive model is developed for the outcome of interest. The participants are then stratified into subgroups based on their predicted baseline risk and a subgroup analysis is performed using these model-defined risk strata rather than single baseline factors. This approach offers the advantage of incorporating all relevant patient characteristics at the same time and offers more patient-centered insights into how treatment effects may differ across the population.

Currently, there is limited research exploring whether the risk of bleeding associated with aspirin use is uniformly distributed among all older adults or whether certain subgroups with different clinical profiles are at a significantly higher risk. The ASPirin in Reducing Events in the Elderly (ASPREE) trial(13), a large-scale randomized trial, gives us an opportunity to explore this knowledge gap. Using a comprehensive dataset of older adults randomized to aspirin or placebo, we apply a risk modeling approach to estimate baseline bleeding risk using a multivariate prediction model. The participants are then stratified by predicted risk to assess whether treatment effects vary significantly between subgroups defined by risk. Our hypothesis is that participants with a higher baseline risk of bleeding will experience greater absolute harm from aspirin therapy compared to those with lower risk. Using machine learning methods, our aim is to provide insights that could help clinicians develop informed evidence-based treatment strategies based on individual patient risk profiles and characteristics.

## Methods

### Data Overview

The ASPREE trial was a randomized, placebo-controlled, double-blind clinical trial that evaluated the effect of enteric-coated 100 mg aspirin on disability-free survival in community-dwelling people aged 70 years and older (or ≥ 65 years for US minorities). The trial methods and main results have been previously published(13,14). Eligible participants had no history of cardiovascular disease, dementia, or physical disability. Key exclusion criteria included elevated risk of bleeding, anemia, prior use of aspirin for secondary prevention, or concurrent use of anticoagulants. Participants taking low-dose NSAIDs or aspirin were eligible if they discontinued use prior to enrollment. The follow-up rates in ASPREE were high, with only 1.2% of the participants (118 assigned to aspirin, 119 to placebo) withdrawing, and 1.5% (139 assigned to aspirin, 157 to placebo) considered lost to follow-up after a median trial follow-up of 4.7 years, having been uncontactable during the final 12 months of the trial(14). All participants provided their informed consent in writing, and the study procedures were approved by the relevant ethics committees.

### Outcome Definition

Our primary outcome was clinically significant bleeding, a composite of hemorrhagic stroke, symptomatic intracranial non-stroke bleeding, or clinically significant extracranial bleeding, as previously detailed(15). Clinically significant extracranial bleeding included bleeding that led to transfusion, hospitalization, prolonged hospitalization, surgery, or death. The investigators of the ASPREE trial required documentation to substantiate clinically significant bleeding. This documentation could include a medical record of the observed bleeding, a credible report of bleeding symptoms, a report of medical personnel, or imaging evidence such as a CT or MRI for intracerebral hemorrhage. Clinically significant bleeding events were adjudicated by an independent clinical events committee blinded to treatment assignment and baseline predictors(15).

### Exposure Definition

Our primary exposure of interest was treatment assignment, with participants randomly assigned in a 1:1 ratio to receive 100 mg of enteric coated aspirin or a corresponding placebo through a computer-generated randomization schedule. For this analysis, treatment exposure was modeled as a time-fixed covariate, based on randomized assignment at baseline (i.e. intention to treat).

### Candidate Predictor Measures

We did not assume any specific relationship between the measures collected by the primary investigators and our main outcome of interest. Instead, we considered all baseline patient characteristics of the ASPREE trial. These predictors were selected based on completeness (<5% missing), clinical relevance, and variation across the sample. Specifically, we excluded non-informative predictors, such as binary variables where >99.9% of values were the same, to avoid redundancy and improve model efficiency. A total of 116 predictor variables were selected through this approach. Various domains are used, such as demographic information (e.g. age, sex, ethnicity), lifestyle factors (e.g. alcohol consumption, smoking history), clinical history (e.g., comorbidities such as cancer, hypertension), physical measures (e.g., BMI, systolic and diastolic blood pressure, waist circumference), laboratory results (e.g., biomarkers such as hemoglobin, creatinine, cholesterol) and medication use (e.g., prescription medications and other agents). Baseline predictors were collected prior to randomization and outcome occurrence, so blinding of predictor assessment was not required. A detailed description of all predictors is provided in the Supplementary Materials, Table D.1. The predictors were not standardized or transformed prior to modeling, since tree-based ensemble methods (RF and RSF) are inherently robust to variable scale and distribution.

For participants with partially missing data (see Supplementary Materials, Table E.1.), we performed multiple imputation using the *missRanger* package in R(16). This method applies random forest-based predictive mean matching to estimate missing values for both continuous and categorical variables and preserves any complex nonlinear relationships and interactions. To avoid potential data leakage, we excluded patient identifiers, treatment assignment, and outcome variables from the imputation step.

### Model Creation

For primary analysis, we used a random forest (RF) model(17) to predict the individual risk of major bleeding using 116 baseline predictors. This model was chosen because of its ability to address several potential challenges with our analysis, particularly overfitting due to having a high-dimensional data set. Using an ensemble of multiple decision trees, RF reduces the risk of overfitting by averaging predictions across trees, and each tree is trained on a bootstrapped sample of the training data. It also ensures that no single predictor or subset of predictors dominates the model by using only a subset of the predictors while creating each tree. In addition, it does not rely on assumptions about the underlying distribution of the data or parameters such as the mean and standard deviation. We fit the model in R using the *ranger* package(18) and listed the model hyperparameters in Supplementary Materials, Table C.1.

Due to the low event rate for our main hemorrhage outcome (3.29%), we trained our models using the full dataset and used out-of-bag (OOB) predictions from each model. OOB prediction is an internal cross-validation method built into ensemble models such as RF and RSF, where each observation is predicted using only the trees that excluded it during the bootstrap sampling stage. This approach allows us to avoid overfitting the model, minimize bias in predicted risks, and maintain statistical power throughout the HTE analyses. To preserve the integrity of the observed outcome distribution, we did not apply any resampling techniques (e.g., oversampling or undersampling), as these methods have been shown to not improve discrimination metrics in internal or external validation and may lead to overestimated risks and miscalibration(19).

### Sensitivity Analysis

To ensure the robustness of our finding and validate the reliability of our primary modeling technique, we performed a sensitivity analysis by also implementing a random survival forest (RSF) model(20). RSF extends the RF methodology by also incorporating time-to-event into the modeling process. Similarly to RF, it also utilizes an ensemble of decision trees, but in this case, each tree is constructed to maximize survival splitting criteria. Survival probability predictions were extracted at a 5-year follow-up from the model, converted to risk probabilities, and then used to generate risk score estimates in a similar fashion to the RF model. The RSF model was fitted in R using the *ranger* package(18) with the hyperparameters listed in Table B.1.

### Model Evaluation

We evaluated the discriminative ability of both models using the area under the receiving operating characteristic curve (AUC). This metric quantifies the model’s ability to distinguish between individuals who experienced a bleeding event and those who did not, with values ranging from 0.5 (no discrimination ability) to 1.0 (perfect discrimination ability). For the RF model, standard (non-time-dependent) AUC was calculated using out-of-bag predictions, and 95% confidence intervals were estimated using the DeLong method(21). For the RSF model, we used time-dependent AUC at 5 years, computed using the inverse probability of censoring weighting (IPCW) approach implemented in the *timeROC* package(22), with 95% confidence intervals computed using the package’s built-in inference procedures. To assess calibration, we computed the Brier score(23) for each model, which quantifies the mean squared difference between predicted probabilities and actual outcomes. Lower Brier values indicate better model calibration, with 0 representing perfect calibration and 1 indicating poor calibration. We also generated calibration plots by grouping participants into deciles based on predicted risk and plotting observed event rates against the mean predicted probability within each decile. To evaluate which baseline variables contributed most to the predicted bleeding risk, we extracted feature importances from both models. Importance was quantified by the total decrease in node impurity (based on the Gini index) attributable to each variable across all trees.

### Heterogeneity of Treatment Effect Analysis

To perform subgroup analysis, we categorized participants into three risk groups (low, intermediate, and high) based on the predicted probability of clinically significant bleeding generated by the RF and RSF models. We defined the cutoff thresholds using the 20th and 80th percentiles of risk probability scores. Risk groups were created separately for each model (RF and RSF) to account for variations in predicted probabilities. The balance of treatment assignment within each stratum was checked to confirm that the risk stratification preserved the original randomization.

We assessed HTE on the relative and absolute scale. On the relative scale, we estimated the cumulative incidence of clinically significant bleeding using a Fine-Gray model fitted with terms for treatment, risk group, and their interaction. This approach was chosen to account for the competing risk of non-hemorrhagic death, which can preclude the observation of a bleeding event and could bias results if treated as noninformative censoring. The Fine-Gray model allows for direct estimation of the subdistribution hazard and cumulative incidence function (CIF) and can provide estimates of the probability of experiencing clinically significant bleeding over time in the presence of competing events. To formally test the presence of interaction effects between treatment and risk group, we assessed the significance of the interaction term using a Type III (Wald) test and a likelihood ratio test (LRT). For absolute treatment effects, we estimated the 5-year CIF for clinically significant bleeding separately for each treatment arm within each risk group. The absolute risk difference (ARD) was calculated as the difference in CIFs between aspirin and placebo. To quantify the uncertainty in estimating ARDs, we used nonparametric bootstrap resampling with 500 iterations to derive 95% confidence intervals. To formally test for HTE in ARDs across risk groups, we used Cochran’s Q test, which evaluates whether the observed differences in ARDs are greater than what would be expected by chance. The Q test was performed using R’s *metafor* package.

## Results

### Participants

The participant flow diagram in Figure 1 provides a detailed overview of the study cohort process. A total of 19,114 participants were enrolled in the ASPREE trial. Using multiple imputation to address missing baseline covariates, we retained all participants in the final analytic cohort. Over a median follow-up of 4.7 years, 626 participants experienced a clinically significant bleeding event, of which approximately 29% were intracranial hemorrhages and 41% were major gastrointestinal bleeding events (upper or lower), with the remaining comprising other clinically significant bleeding events.

**Figure 1.**
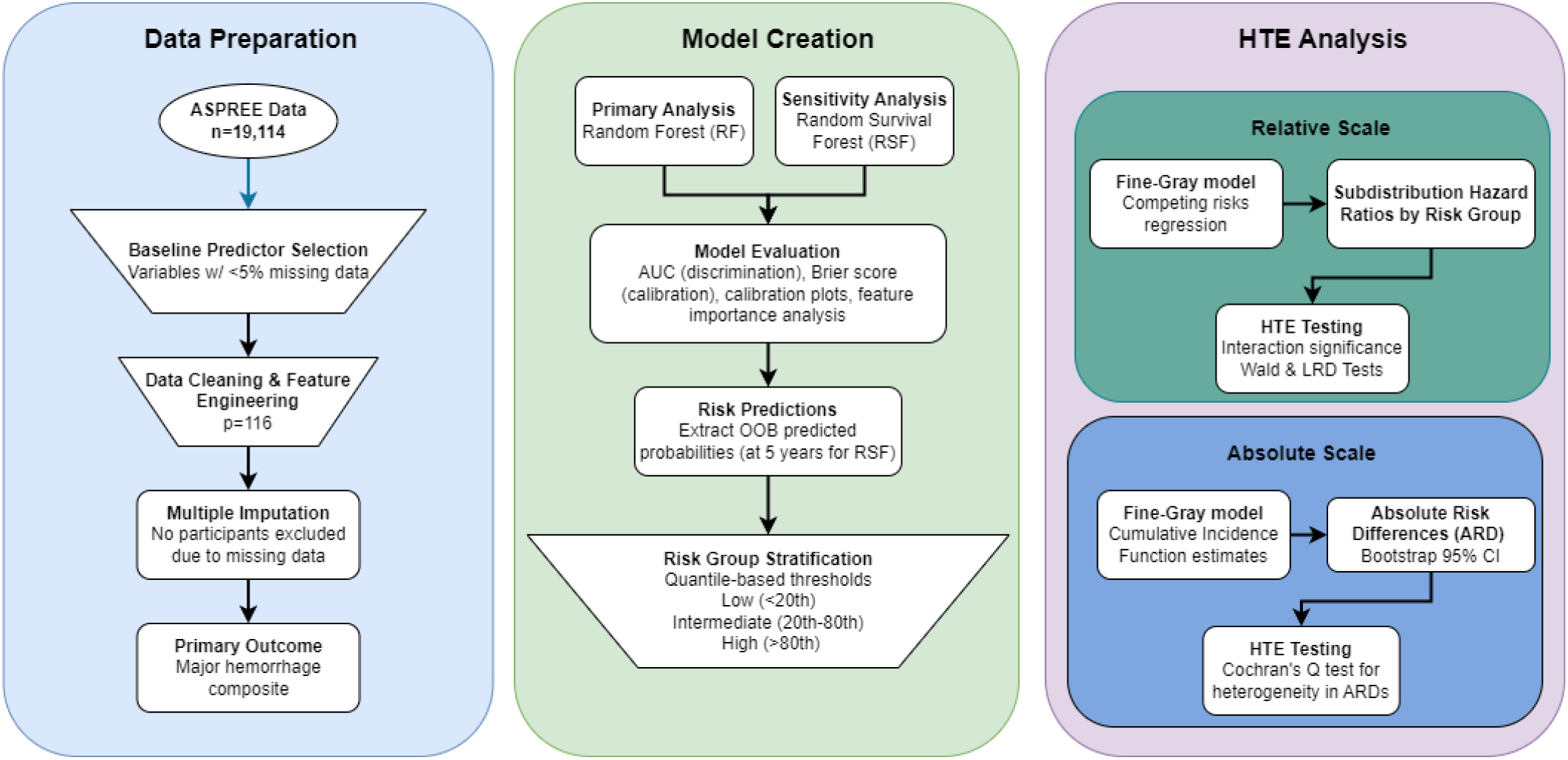
Participant flow diagram and methodology overview.

### Model Performance

The OOB AUC for the RF model was 0.65 (95% CI: 0.63–0.67), and for the RSF model was 0.65 (95% CI: 0.63–0.68) (Supplementary Materials, Figures C.1 and B.1). Both models showed moderate discriminative ability in predicting individuals who would experience a clinically significant bleeding event versus those who would not. Both models also had similar calibration performance (RF: Brier = 0.032, 95% CI: 0.029-0.034; RSF: Brier = 0.032, 95% CI: 0.029-0.034). The calibration plots for both the RF (Supplementary Materials, Figure C.2) and the RSF (Supplementary Materials, Figure B.2) models generally showed good agreement between the predicted and observed risks within deciles, with minor deviations towards the highest predicted risk levels.

### Validation of Risk-Based Subgroups

We examined the distribution of the predicted clinically significant bleeding risk across the entire study population. The distribution was right-skewed, with most participants assigned lower predicted risks and a smaller proportion at higher risk (Figure 2.a). We also compared the predicted risk distributions separately by treatment arm to ensure balance in risk modeling. We found no significant variation for the predicted risk between treatment arms (Figure 2.b). To define clinically relevant subgroups for HTE analysis, we stratified participants into low, intermediate, and high-risk groups using quantile-based thresholds. We used the 20th percentile and 80th percentile of the predicted risk distribution to define the cutoffs for the three categories. Participants with predicted risk probabilities below the 20th percentile were assigned to the low-risk group, those between the 20th and 80th percentiles to the intermediate-risk group, and those above the 80th percentile to the high-risk group. The distribution of predicted risk probabilities was nearly identical between aspirin and placebo arms.

**Figure 2.**
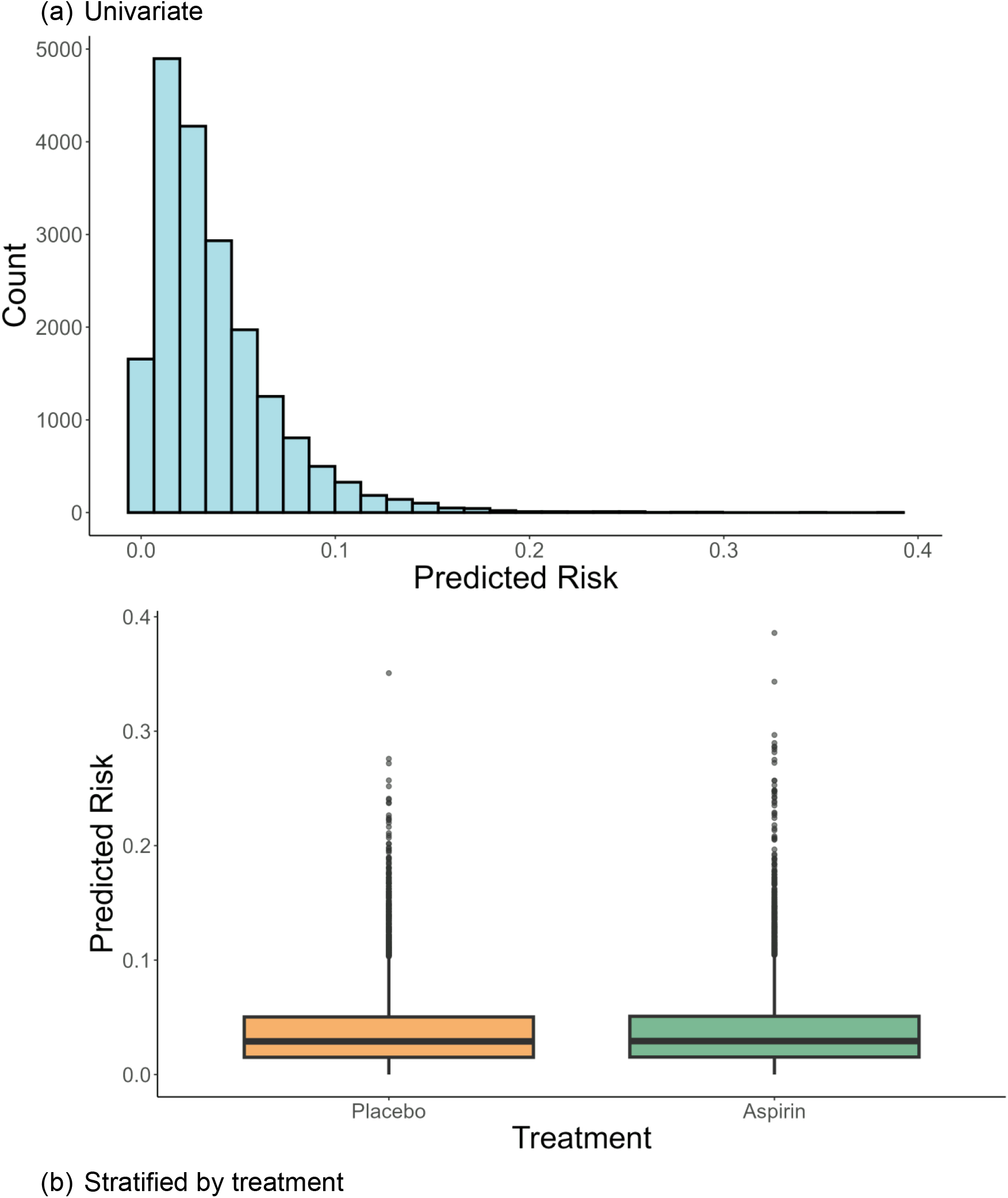
Histograms of predicted bleeding risk scores based on the RF model. Panel (a) shows the full risk distribution across all participants. Panel (b) shows the risk distribution stratified by treatment assignment.

Figure 3 shows the cumulative incidence of clinically significant bleeding over time, stratified by predicted risk group. There is clear and progressive separation between groups, with the highest-risk group having the highest cumulative incidence, followed by the intermediate and then low-risk group. At approximately 5 years of follow-up, the estimated cumulative incidence of clinically significant bleeding was 1.4% in the low-risk group, 3.2% in the intermediate-risk group, and 6.5% in the high-risk group. These results support the discriminative capacity of the risk stratification model and the clinical relevance of assigning individuals to risk-based categories. Median time to clinically significant bleeding for clinically significant bleeding was not reached in any risk group during the follow-up period due to the relatively low event rate.

**Figure 3.**
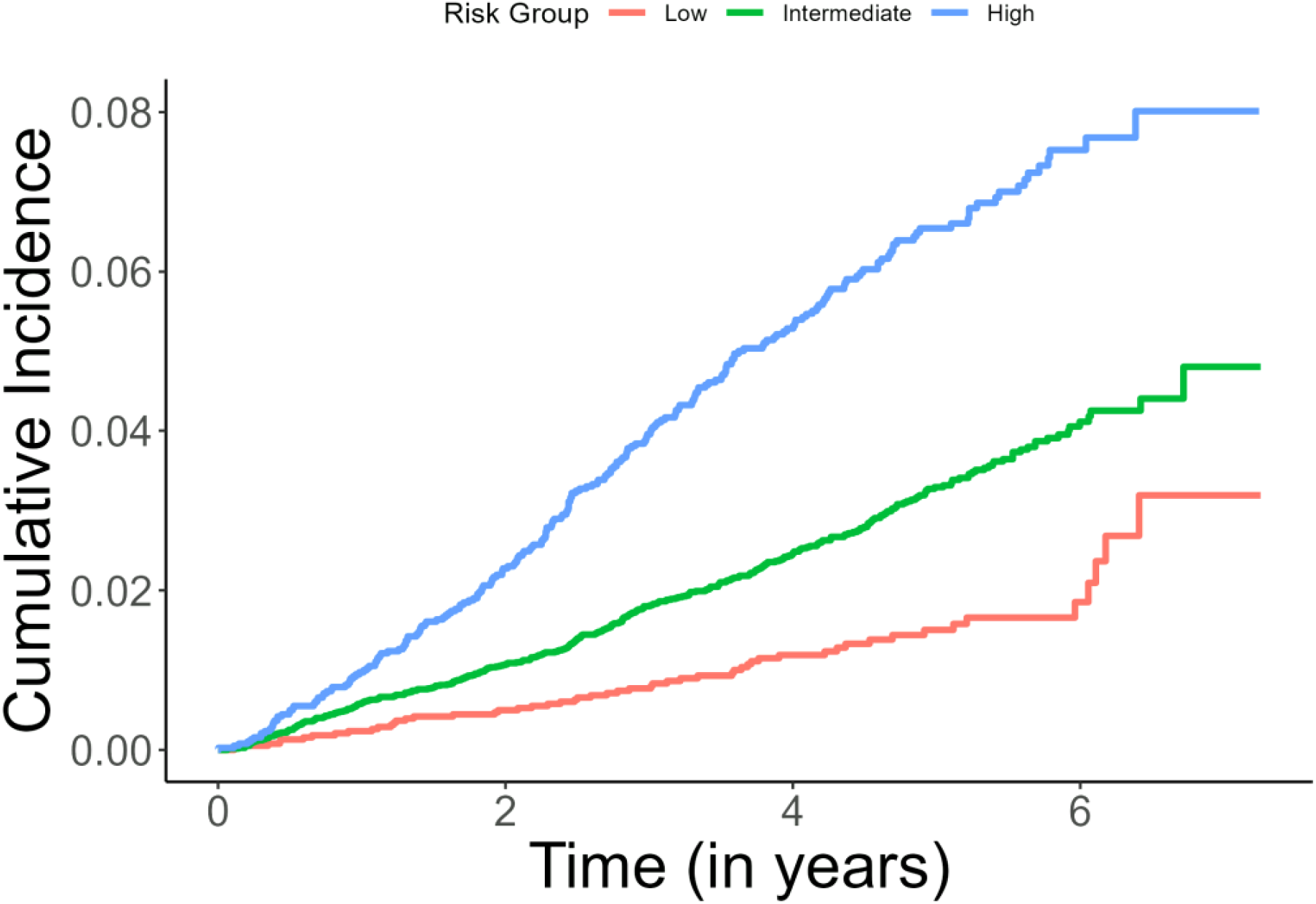
Cumulative incidence of clinically significant bleeding stratified by risk group.

### Heterogeneity of Treatment Effect Results

We fitted a Fine-Gray competing risks model with terms for treatment, risk group, and their interaction to account for the competing risk of non-hemorrhagic death and to formally test for heterogeneity of treatment effects. On the relative scale, the interaction between treatment assignment and the risk group was statistically significant (p = 0.027), which suggests that the relative effect of treatment is not uniform between all participants, and the magnitude of the effect associated with aspirin differs significantly depending on the predicted risk of bleeding of an individual. Subdistribution hazard ratios (SHRs) for aspirin versus placebo were highest in the low-risk group (HR = 2.26, 95% CI: 1.27 to 4.01), followed by the intermediate-risk group (HR = 1.50, 95% CI: 1.21 to 1.86), and lowest in the high-risk group (HR = 1.07, 95% CI: 0.82 to 1.38). These results suggest that the relative harm associated with aspirin is greatest among those at a lower baseline risk and diminishes as baseline bleeding risk increases. The trend is visually represented in Figure 4 and summarized in Table 2. This same pattern of relative treatment effect and interaction significance was observed when stratifying participants by RSF-derived risk groups (Supplementary Materials, Table B.3 and Figure B.5), maintaining consistency across both modeling approaches.

**Table 1.** Cumulative incidence function values by group at evaluation times closest to 5 years, stratified by risk group. The main outcome of interest is clinically significant bleeding.

| Group | Eval_Time | CIF | CIF (%) |
| --- | --- | --- | --- |
| Low | 4.914 | 0.014 | 1.4% |
| Intermediate | 4.986 | 0.033 | 3.3% |
| High | 5.101 | 0.065 | 6.5% |

**Table 2.** Subdistribution hazard ratios and 95% confidence intervals for aspirin versus placebo, stratified by predicted baseline bleeding risk.

| Risk Group | HR | Lower CI | Upper CI |
| --- | --- | --- | --- |
| Low | 2.257 | 1.272 | 4.005 |
| Intermediate | 1.501 | 1.208 | 1.864 |
| High | 1.067 | 0.825 | 1.380 |

**Figure 4.**
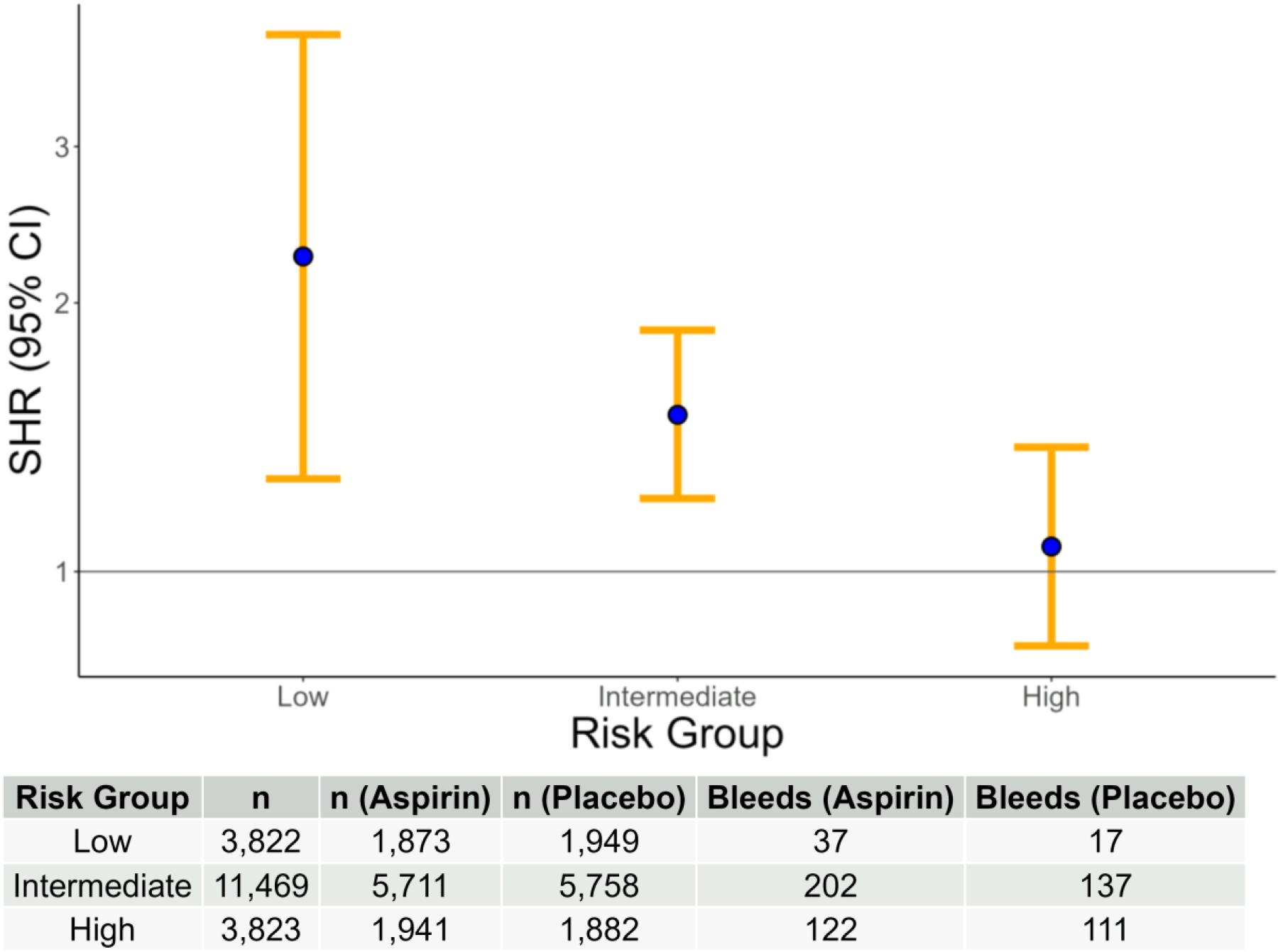
Subdistribution hazard ratios with 95% confidence intervals for clinically significant bleeding comparing aspirin to placebo, stratified by predicted bleeding risk groups derived from the Random Forest model. The risk group-specific counts and number of bleeding events are shown below the plot.

We also evaluated HTE on the absolute scale by estimating 5-year cumulative incidence functions from the Fine-Gray model and calculating absolute risk differences between treatment groups within each risk stratum. As shown in Table 3, the 5-year cumulative incidence of clinically significant bleeding was 1.06% in the low-risk group for aspirin and 0.59% for placebo, with an ARD of 1.17% (95% CI: 0.37% to 1.95%). In the intermediate-risk group, the ARD was 1.32% (95% CI: 0.63% to 2.01%), while in the high-risk group, the difference was smaller and not statistically significant (ARD = 0.40%, 95% CI: –1.29% to 1.96%). These findings are visualized in Figure 5. Although the point estimates for the lower and intermediate risk groups suggest greater absolute risk increases, the formal test for heterogeneity across risk strata using Cochran’s Q test was not statistically significant (Q = 1.02, p = 0.60), so we cannot conclude that absolute risk differences varied meaningfully across risk groups. The findings were consistent with those obtained using RSF-based risk groups (Supplementary Materials, Tables B.5 and B.6).

**Table 3.** Cumulative incidence of clinically significant bleeding at 5 years for aspirin and placebo groups, stratified by predicted baseline bleeding risk. Positive ARDs indicate increased bleeding risk with aspirin, and the number needed to harm (NNH) reflects the number of patients who would need to be treated with aspirin for one additional major bleeding event to occur.

| Risk Group | CIF (Aspirin) | CIF (Placebo) | ARD | Upper CI | Lower CI | NNH |
| --- | --- | --- | --- | --- | --- | --- |
| Low | 0.0106 | 0.0059 | 0.0117 | 0.0037 | 0.0195 | 86 |
| Intermediate | 0.0203 | 0.0125 | 0.0132 | 0.0062 | 0.0201 | 76 |
| High | 0.0338 | 0.0318 | 0.0040 | -0.0129 | 0.0196 | 250 |

**Figure 5.**
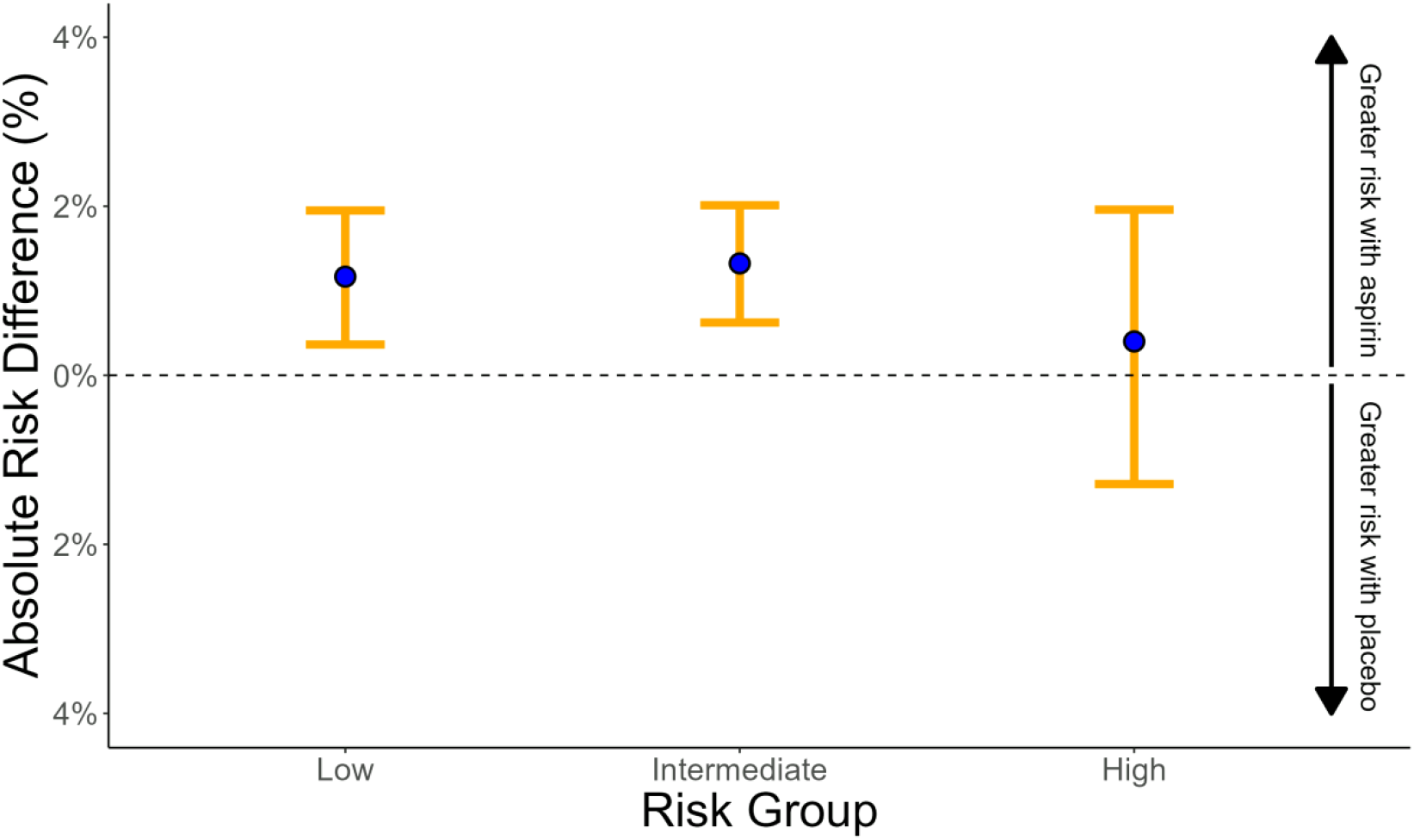
Absolute risk differences at 5 years comparing aspirin to placebo, stratified by predicted baseline bleeding risk. Positive values indicate greater bleeding risk with aspirin, while negative values would indicate greater risk with placebo.

### Clinical Profiles Across Risk Strata

We also assessed the importance of the features used in both the RF and RSF models to predict bleeding risk. The top 30 predictors for the RF model are shown in Supplementary Materials, Figure C.2 and include age, SF-12 mental and physical scores, estimated glomerular filtration rate (eGFR), weight, average gait speed, and other measures of physical function and cardiometabolic health. Similarly, the top features for the RSF model, shown in Supplementary Materials, Figure B.2, included many of the same variables, such as creatinine, glucose, antithrombotic use, diastolic blood pressure, and mean heart rate. Both of our models consistently identified physiological and functional status indicators as key drivers of bleeding risk.

To understand the clinical factors that drive HTE, we examined the distribution of the top random forest predictors across risk strata (Supplementary Materials, Table A.1). Our analysis revealed distinct clinical profiles by risk group that can potentially explain the observed treatment effect heterogeneity. In terms of physical function and frailty markers, significant differences were observed across risk groups. Mean age increased progressively from 73.21 years in the low-risk group to 77.82 years in the high-risk group (p<0.001). Physical functioning scores (SF-12 Physical) declined substantially from 51.37 to 43.94 (p<0.001), while gait speed slowed from 2.84 to 3.67 seconds per 3 meters (p<0.001). Cognitive performance, measured by HVLT total score, similarly declined from 32.60 to 27.45 across risk groups (p<0.001). Kidney function also deteriorated from low- to high-risk groups, with mean eGFR declining from 75.66 to 69.56 mL/min/1.73m^2^ and serum creatinine rising from 75.98 to 84.30 μmol/L (both p<0.001). Another notable difference across risk groups is concurrent medication exposure. Antithrombotic agent use showed a six-fold increase from low- to high-risk groups (6.1% vs 36.6%, p<0.001), while cardiac therapy use increased 21-fold (0.5% vs 10.6%, p<0.001). Regular aspirin use prior to enrollment also increased substantially across risk groups (10.9% vs 16.5%, p<0.001).

## Discussion

In this study, we used machine learning models to investigate whether the effect of aspirin on clinically significant bleeding risk in older adults varies by predicted risk. Our hypothesis was that participants with a higher baseline risk of clinically significant bleeding would experience greater absolute harm from aspirin therapy compared to those at lower risk. However, our findings did not support this expectation. Instead, we observed the opposite pattern, where participants in both the low-risk and intermediate-risk groups experienced statistically significant increases in bleeding risk when treated with aspirin, while the high-risk group showed no significant increase. Using both RF and RSF approaches, we found statistically significant evidence of treatment effect heterogeneity between risk categories on the relative scale. In both models, the interaction between the assignment of treatment and the baseline bleeding risk group was significant, which is evidence that the relative treatment effect of aspirin is not uniform between individuals. Specifically, both low-risk and intermediate-risk participants showed statistically significant increases in bleeding risk with aspirin (low-risk: RF SHR = 2.26, 95% CI: 1.27 to 4.01; intermediate-risk: RF SHR = 1.50, 95% CI: 1.21 to 1.86), while the high-risk group showed no significant increase (RF SHR = 1.07, 95% CI: 0.82 to 1.38). On the absolute scale, we observed increases in the 5-year cumulative incidence of clinically significant bleeding among aspirin users across all of the risk strata, but the formal test for heterogeneity in absolute risk differences between risk groups was not statistically significant. These findings support the presence of heterogeneity of treatment effect on the relative scale, with consistent patterns observed using both RF- and RSF-derived risk stratifications.

While the participants in the high-risk group had the highest absolute rates of clinically significant bleeding, the counterintuitive finding was that participants in the lower baseline risk groups (both low- and intermediate-risk) experienced statistically significant increases in bleeding risk with aspirin, whereas those at highest baseline risk showed no significant increase. This challenges conventional clinical assumptions and we believe may warrant further examination. The baseline characteristic differences identified through risk stratification and study treatment allocation (Supplementary Materials, Tables A.1. and F.1.) provide several potential mechanisms which may drive the observed treatment effect heterogeneity. The most likely explanation may relate to differences in medication exposure history. In the low- and intermediate-risk groups, 88.4% and 78.9% of the patients had never used antithrombotic medications, compared to only 60.5% in the high-risk group. This would suggest that the low- and intermediate-risk groups had normal, unaltered blood clotting systems. When given aspirin, their bodies may react more strongly because they were not exposed to antiplatelet drugs before, which may have unmasked their underlying propensity for bleeding.

This suggests that individuals in the low- and intermediate-risk groups may have had normal, uncompromised hemostatic systems prior to the trial. Upon exposure to aspirin—likely their first antiplatelet agent—previously unrecognized susceptibility to bleeding may have been unmasked, resulting in a disproportionately higher bleeding risk compared to those in the high-risk group, who may have had prior exposure or other mitigating factors influencing baseline risk.

On the other hand, people in the high-risk group may represent a population with a survivorship effect, where those who experienced earlier adverse bleeding outcomes or tolerance issues to antithrombotic therapy were no longer being treated or eligible for enrollment. Additionally, high-risk participants had a substantially higher use of cardiac therapies (10.6% vs 0.5% and 2.7%) and more frequent regular aspirin use prior to enrollment (16.5% vs 6.8% and 10.5%), which likely reflects ongoing cardiovascular care. This experience may have included dose adjustments, monitoring, and protective treatments (such as proton pump inhibitors), which would create conditions that reduce aspirin’s added bleeding risk compared to those who were medication naïve.

To our knowledge, no other analysis has examined the heterogeneity modeling of treatment effects using the Predictive Approaches to Heterogeneous Treatment Effects (PATH) (12) framework with outcome risk prediction derived from random forest models to evaluate the randomization of aspirin versus placebo in older adults. This analysis is particularly novel because it focuses on a population without a clinical indication for aspirin therapy and without known contraindications to its use.

This study has some limitations. First, while our machine learning ensemble models were able to identify key predictors of major bleeding and allowed for risk stratification, they are designed for prediction rather than for estimating individual-level causal effects. Second, the ability of the models to predict the risk of bleeding may have been constrained by the variables available in the data set. Some clinically relevant predictors of bleeding may not have been collected, because major bleeding was a secondary outcome in the ASPREE trial, which was primarily designed to evaluate disability-free survival. Third, our analysis treated clinically significant bleeding as a composite outcome that comprises different types of bleeding, including intracranial hemorrhages (29% of events), gastrointestinal bleeding (41% of events), and other clinically significant bleeding events (30% of events). Each of these bleeding subtypes may have a distinct risk profiles and mechanisms, and potentially different heterogeneity patterns in response to aspirin therapy. Lastly, the generalizability of our findings may be limited to populations similar to those in the ASPREE cohort, specifically older adults without a clinical indication for aspirin therapy, so caution should be taken when extrapolating the results to other groups.

This study also has several key strengths. First, it benefits from careful adjudication of clinically significant bleeding events in a large cohort of healthy older adults. Second, the ASPREE trial had strong follow-up procedures and high participant retention rates. Third, we used two independent ensemble modeling techniques (random forest and random survival forest) and obtained similar results between both methods. Fourth, we used out-of-bag estimation to assess model performance and minimize overfitting problems. Fifth, we accounted for competing risks using Fine-Gray subdistribution hazard models, which provide more accurate estimates of cumulative incidence in the presence of competing events such as non-bleeding death. Finally, we used a comprehensive set of more than 100 baseline predictors to develop a bleeding risk prediction model that was independent of treatment assignment.

## Conclusion

In analyzing data from more than 19,000 older adults randomized to aspirin or placebo, we found evidence of heterogeneity in the risk of aspirin-associated clinically significant bleeding, with the magnitude of harm differing between baseline risk groups. Formal statistical tests of interaction reached conventional thresholds for significance, particularly on the relative scale, where treatment effect heterogeneity was most apparent. Corroborating evidence was also observed on the absolute scale, though tests for heterogeneity in absolute risk differences were not statistically significant. Our results indicate that the risk of major bleeding associated with aspirin exposure in older adults is not uniform across the population and that specific subgroups can be identified that may be at a greater risk of bleeding events when starting aspirin therapy. If confirmed, these insights could aid in clinical decision-making by both clinicians and older adults, by identifying individuals who may require additional precautions when considering self-use of aspirin. Although current guidelines generally advise against the use of aspirin in older adults with asymptomatic aging, further research on risk-adapted approaches to aspirin therapy may be warranted.

## Data Availability

For access to the ASPirin in Reducing Events in the Elderly (ASPREE) project data, visit ams.aspree.org. Code for this project, including final hyperparameters and model implementation, is available at: https://anonymous.4open.science/r/P428_MajorHem/README.md.

## Supplementary Materials

The supplementary materials file contains six appendices. Appendix A provides a summary table of the baseline variables by low, medium, and high bleeding risk group designation. Appendix B contains the detailed figures and tables for risk group determination and examination of heterogeneity of treatment effect of aspirin versus placebo when random survival forest models were utilized instead of random forest models. Appendix C contains tables and figures for extra details of the random forest models utilized in the main paper. Appendix D provides a descriptive listing of all predictor variables utilized in the models for risk group determination. Appendix E summarizes the number of participants with missing values for given predictor variables at baseline which then were imputed. Appendix F provides a table of the baseline characteristics by assignment to study treatment in the ASPREE.

## Acknowledgements

The authors recognize the significant contributions made by the research participants, staff, and investigators for the ASPirin in Reducing Events in the Elderly clinical trial (Clinical trial registry number: NCT01038583).

