## Supplemental Material for "Heterogeneity of Treatment Effect of Aspirin and Clinically Significant Bleeding in Older Adults"

#### TABLE OF CONTENTS

**Page**

1. **Appendix A: Summary Table of Baseline Variables by Risk Group** **3**
   a. **Table A.1:** Baseline characteristics of the top 30 most important predictors from the Random Forest model, stratified by predicted bleeding risk group. P-values were calculated using ANOVA for continuous variables and Fisher's exact or chi-square test for categorical variables, as appropriate 3-4
2. **Appendix B: Random Survival Forest Sensitivity Analysis Results** **5**
   a. **Table B.1:** Model configuration and performance metrics for the random survival forest (RSF) trained to predict clinically significant bleeding 5
   b. **Figure B.1:** Receiver operating characteristic (ROC) curve at 5 years for the random survival forest model predicting clinically significant bleeding 6
   c. **Figure B.2**: Calibration plot for the Random Survival Forest model predicting clinically significant bleeding risk. The plot compares observed event rates to predicted probabilities across deciles of predicted risk 7
   d. **Figure B.3:** Top predictors of clinically significant bleeding identified by the RSF model using impurity-based variable importance. Higher values indicate greater contribution to the model's predictive accuracy. 8
   e. **Figure B.4:** Distribution of predicted bleeding risk scores based on the RSF model. Panel (a) shows the full risk distribution across all participants. Panel (b) shows the risk distribution stratified by treatment assignment. 8-9
   f. **Table B.2:** Summary statistics for predicted 5-year bleeding risk scores from the random survival forest model, stratified by RSF-based risk groups. 10
   g. **Table B.3:** Results from a Fine-Gray competing risks regression model evaluating the effect of aspirin versus placebo across RSF-derived bleeding risk groups. The model includes interaction terms to assess heterogeneity in treatment effect by risk group 11
   h. **Figure B.5:** Hazard ratios and 95% confidence intervals for aspirin versus placebo within each RSF-derived bleeding risk group, estimated using a Fine-Gray competing risks model. Below the plot, the table summarizes group sizes and the number of clinically significant bleeding events by treatment arm. 11
   i. **Table B.3:** Subdistribution hazard ratios and 95% confidence intervals for aspirin versus placebo within each RSF-based bleeding risk group, derived from a Fine-Gray competing risks model.
   j. **Table B.4:** Results from likelihood ratio and Wald tests evaluating the significance of treatment, RSF-derived bleeding risk groups, and their interaction in the Fine-Gray competing risks model. 12
   k. **Table B.5:** Cumulative incidence of clinically significant bleeding at 5 years for aspirin and placebo groups, stratified by RSF-based baseline bleeding risk. Positive ARDs indicate increased bleeding risk with aspirin, and the number needed to harm (NNH) reflects the number of patients who would need to be treated with aspirin for one additional major bleeding event to occur. 12

l. **Table B.6:** Results from a Q-test evaluating heterogeneity in ARDs between aspirin and placebo at 5 years across RSF-derived risk groups. 13
m. **Figure B.6:** Cumulative incidence functions for clinically significant bleeding over time, stratified by RSF-based risk group. 13
n. **Table B.7:** Cumulative incidence function values by group at evaluation times closest to 5 years, stratified by RSF-based risk group. 14
o. **Figure B.7:** Stratified cumulative incidence curves for clinically significant bleeding by treatment arm (aspirin vs. placebo) within each RSF-derived bleeding risk group. 14
p. **Table B.8:** Cumulative incidence function estimates at approximately 5 years, stratified by treatment group, RSF-derived bleeding risk group, and event type. 15

**3. Appendix C: Random Forest Additional Outputs** **16**

a. **Table C.1:** Summary of Random Forest model parameters used to estimate the probability of clinically significant bleeding. 16
b. **Figure C.1:** Receiver operating characteristic (ROC) curve at 5 years for the random forest model predicting clinically significant bleeding 17
**c.** **Figure C.2:** Calibration plot for the Random Forest model predicting clinically significant bleeding risk. The plot compares observed event rates to predicted probabilities across deciles of predicted risk. 18
d. **Figure C.3:** Top predictors of clinically significant bleeding identified by the RF model using impurity-based variable importance. Higher values indicate greater contribution to the model's predictive accuracy. 19
e. **Table C.2:** Results from a Fine-Gray competing risks regression model evaluating the effect of aspirin versus placebo across Random Forest–derived bleeding risk groups. The model includes interaction terms to assess heterogeneity in treatment effect by risk group. 20
f. **Table C.3:** Results from likelihood ratio and Wald tests evaluating the significance of treatment, RF-derived bleeding risk groups, and their interaction in the Fine-Gray competing risks model. 20
g. **Table C.4**: Results from a Q-test evaluating heterogeneity in absolute risk differences between aspirin and placebo at 5 years across Random Forest–derived bleeding risk groups. 21
h. **Figure C.4:** Stratified cumulative incidence curves for clinically significant bleeding by treatment
i. arm (aspirin vs. placebo) within each RF-derived bleeding risk group. 21
j. **Table C.5:** Cumulative incidence function estimates at approximately 5 years, stratified by treatment group, RF-derived bleeding risk group, and event type. 22

4. **Appendix D: Predictor Variables and Descriptions** **23**

a. **Table D.1:** List of predictor variables and descriptions used in model development. 23-30

5. **Appendix E: Pre-Imputation Missing Value Summary** **31**

a. **Table E.1:** Number of missing values for each feature prior to imputation. Many features with identical missingness (n = 868) were medication-related and are grouped under a single row labeled Medication variables. 31-33

**6. Appendix F: Baseline Characteristics of ASPREE Participants by Treatment Group** **34**

a. **Table F.1:** Summary of demographic, clinical, lifestyle, and medication-related baseline characteristics of ASPREE participants, stratified by treatment assignment. 34-41

### Appendix A: Summary Table of Baseline Variables by Risk Group

**Table A.1:** Baseline characteristics of the top 30 most important predictors from the Random Forest model, stratified by predicted bleeding risk group. P-values were calculated using ANOVA for continuous variables and Fisher's exact or chi-square test for categorical variables, as appropriate.

| | Stratified by RF Risk Groups | | | |
|---|---|---|---|---|
| | Low | Intermediate | High | p |
| n | 3,822 | 11,469 | 3,823 | |
| Age (mean (SD)) | 73.21 (2.78) | 74.84 (4.10) | 77.82 (5.84) | <0.001 |
| eGFR_CKD (mean (SD)) | 75.66 (10.80) | 73.07 (13.47) | 69.56 (17.05) | <0.001 |
| Weight (mean (SD)) | 76.38 (11.86) | 77.15 (14.43) | 77.08 (18.82) | 0.020 |
| SF12_Mental (mean (SD)) | 56.64 (4.99) | 55.80 (6.80) | 54.31 (9.36) | <0.001 |
| SF12_Physical (mean (SD)) | 51.37 (6.30) | 48.79 (8.26) | 43.94 (10.51) | <0.001 |
| AvgGaitSpd (mean (SD)) | 2.84 (0.52) | 3.08 (0.81) | 3.67 (1.38) | <0.001 |
| Creat_molL (mean (SD)) | 75.98 (14.22) | 80.02 (18.11) | 84.30 (26.12) | <0.001 |
| AvgDominantGrpStr (mean (SD)) | 27.65 (8.96) | 27.33 (9.88) | 24.74 (11.08) | <0.001 |
| HeartRateMean (mean (SD)) | 69.91 (9.00) | 70.58 (10.57) | 71.96 (12.54) | <0.001 |
| HVLT_Total (mean (SD)) | 32.60 (6.95) | 30.32 (7.80) | 27.45 (8.50) | <0.001 |
| Hemoglobin (mean (SD)) | 14.24 (1.04) | 14.22 (1.20) | 13.92 (1.40) | <0.001 |
| SDMT_Score (mean (SD)) | 40.80 (8.52) | 37.07 (9.80) | 31.65 (10.56) | <0.001 |
| SBPMean (mean (SD)) | 137.06 (14.19) | 139.17 (16.39) | 141.37 (18.59) | <0.001 |
| AbdCirc (mean (SD)) | 95.81 (10.42) | 97.25 (12.39) | 98.20 (15.96) | <0.001 |
| TotalCholesterol_mmolL (mean (SD)) | 5.37 (0.79) | 5.22 (0.97) | 5.15 (1.17) | <0.001 |
| Height (mean (SD)) | 1.65 (0.08) | 1.66 (0.09) | 1.65 (0.10) | <0.001 |
| Tg_mmolL (mean (SD)) | 1.25 (0.50) | 1.32 (0.63) | 1.41 (0.85) | <0.001 |
| Glucose_mmolL (mean (SD)) | 5.33 (0.62) | 5.48 (0.97) | 5.65 (1.49) | <0.001 |
| LDL_mmolL (mean (SD)) | 3.17 (0.72) | 3.04 (0.87) | 2.93 (1.01) | <0.001 |
| DBPMean (mean (SD)) | 77.18 (8.49) | 77.39 (9.91) | 76.96 (11.45) | 0.055 |
| HDL_mmolL (mean (SD)) | 1.62 (0.39) | 1.57 (0.45) | 1.58 (0.54) | <0.001 |
| COWAT_Score (mean (SD)) | 12.83 (4.28) | 12.08 (4.51) | 11.39 (4.87) | <0.001 |
| X3MS_Overall (mean (SD)) | 94.87 (3.60) | 93.50 (4.49) | 91.74 (5.34) | <0.001 |
| HVLT_Slope (mean (SD)) | 2.46 (1.95) | 2.14 (2.25) | 1.82 (2.44) | <0.001 |
| CESD_Overall (mean (SD)) | 2.48 (2.44) | 3.07 (3.14) | 4.26 (4.15) | <0.001 |
| Education (%) | | | | <0.001 |
| <9 yrs | 455 (11.9) | 1,778 (15.5) | 769 (20.1) | |
| 9-11 yrs | 1,132 (29.6) | 3,409 (29.7) | 1,093 (28.6) | |
| 12 yrs | 451 (11.8) | 1,368 (11.9) | 500 (13.1) | |
| 13-15 yrs | 683 (17.9) | 1,929 (16.8) | 643 (16.8) | |
| 16 yrs | 415 (10.9) | 1,042 ( 9.1) | 309 ( 8.1) | |
| 17-21 yrs | 686 (17.9) | 1,943 (16.9) | 509 (13.3) | |

| | | | | |
|---|---|---|---|---|
| BMI (%) | | | | <0.001 |
| Underweight | 0 ( 0.0) | 20 ( 0.2) | 83 ( 2.2) | |
| Normal | 804 (21.0) | 2,990 (26.1) | 1,086 (28.4) | |
| Overweight | 2,014 (52.7) | 5,078 (44.3) | 1,420 (37.1) | |
| Obese | 1,004 (26.3) | 3,381 (29.5) | 1,234 (32.3) | |
| AntithromboticAgents (%) | | | | <0.001 |
| No | 3,380 (88.4) | 9,052 (78.9) | 2,312 (60.5) | |
| Yes | 234 ( 6.1) | 1,868 (16.3) | 1,400 (36.6) | |
| Not recorded | 208 ( 5.4) | 549 ( 4.8) | 111 ( 2.9) | |
| Smoking (%) | | | | <0.001 |
| Current | 49 ( 1.3) | 419 ( 3.7) | 267 ( 7.0) | |
| Former | 1,480 (38.7) | 4,712 (41.1) | 1,607 (42.0) | |
| Never | 2,293 (60.0) | 6,338 (55.3) | 1,949 (51.0) | |
| CardiacTherapy (%) | | | | <0.001 |
| No | 3,594 (94.0) | 10,611 (92.5) | 3,307 (86.5) | |
| Yes | 20 ( 0.5) | 309 ( 2.7) | 405 (10.6) | |
| Not recorded | 208 ( 5.4) | 549 ( 4.8) | 111 ( 2.9) | |
| RegAspirin = Yes (%) | 260 ( 6.8) | 1,204 (10.5) | 631 (16.5) | <0.001 |
| Treatment = Aspirin (%) | 1,873 (49.0) | 5,711 (49.8) | 1,941 (50.8) | 0.301 |
| MajorHem = 1 (%) | 54 ( 1.4) | 339 ( 3.0) | 233 ( 6.1) | <0.001 |

### Appendix B: RSF Sensitivity Analysis Results

**Table B.1:** Model configuration and performance metrics for the random survival forest (RSF) trained to predict clinically significant bleeding.

| Parameter | Value |
|---|---|
| Model Type | Survival |
| Number of Trees | 500 |
| Sample Size | 19,114 |
| Number of Independent Variables | 116 |
| mtry | 11 |
| Target Node Size | 3 |
| Variable Importance Mode | Impurity |
| Split Rule | Logrank |
| Number of Unique Death Times | 542 |
| OOB Prediction Error (1 - C) | 0.3450804 |

**Figure B.1:** Receiver operating characteristic (ROC) curve at 5 years for the random survival forest model predicting clinically significant bleeding.

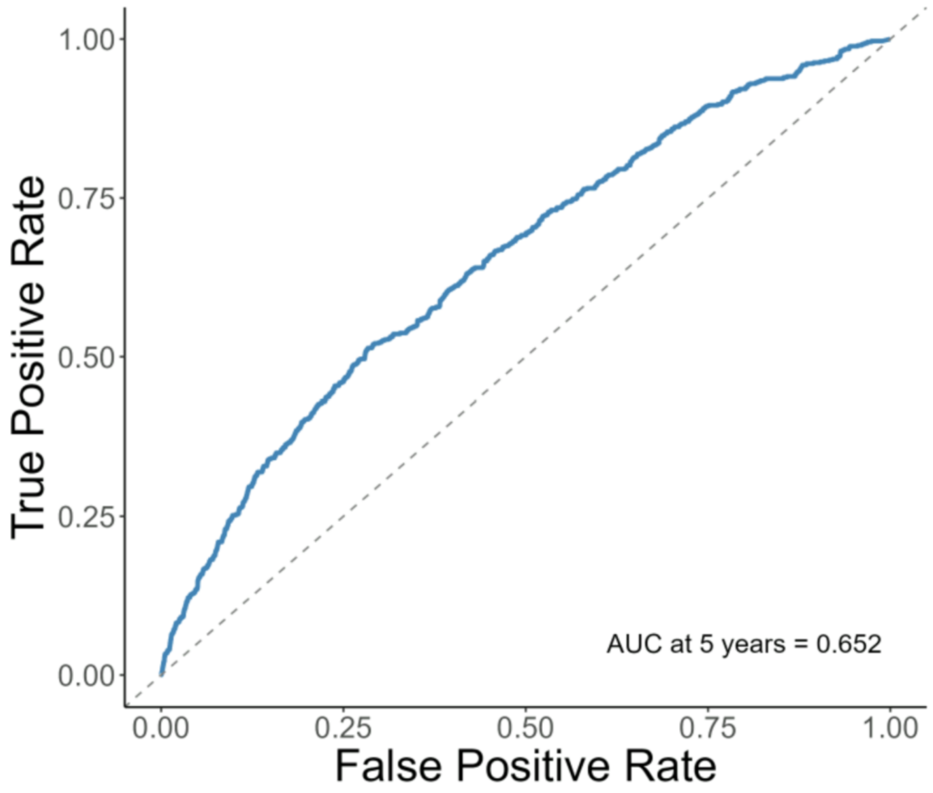

**Figure B.2:** Calibration plot for the Random Survival Forest model predicting clinically significant bleeding risk. The plot compares observed event rates to predicted probabilities across deciles of predicted risk.

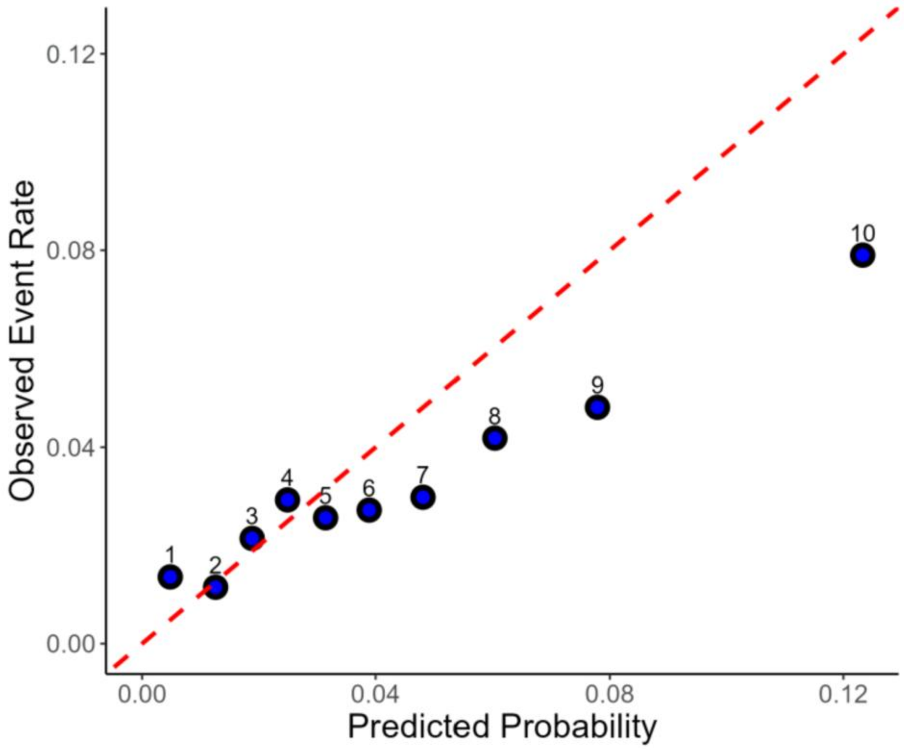

**Figure B.3:** Top predictors of clinically significant bleeding identified by the RSF model using impurity-based variable importance. Higher values indicate greater contribution to the model's predictive accuracy.

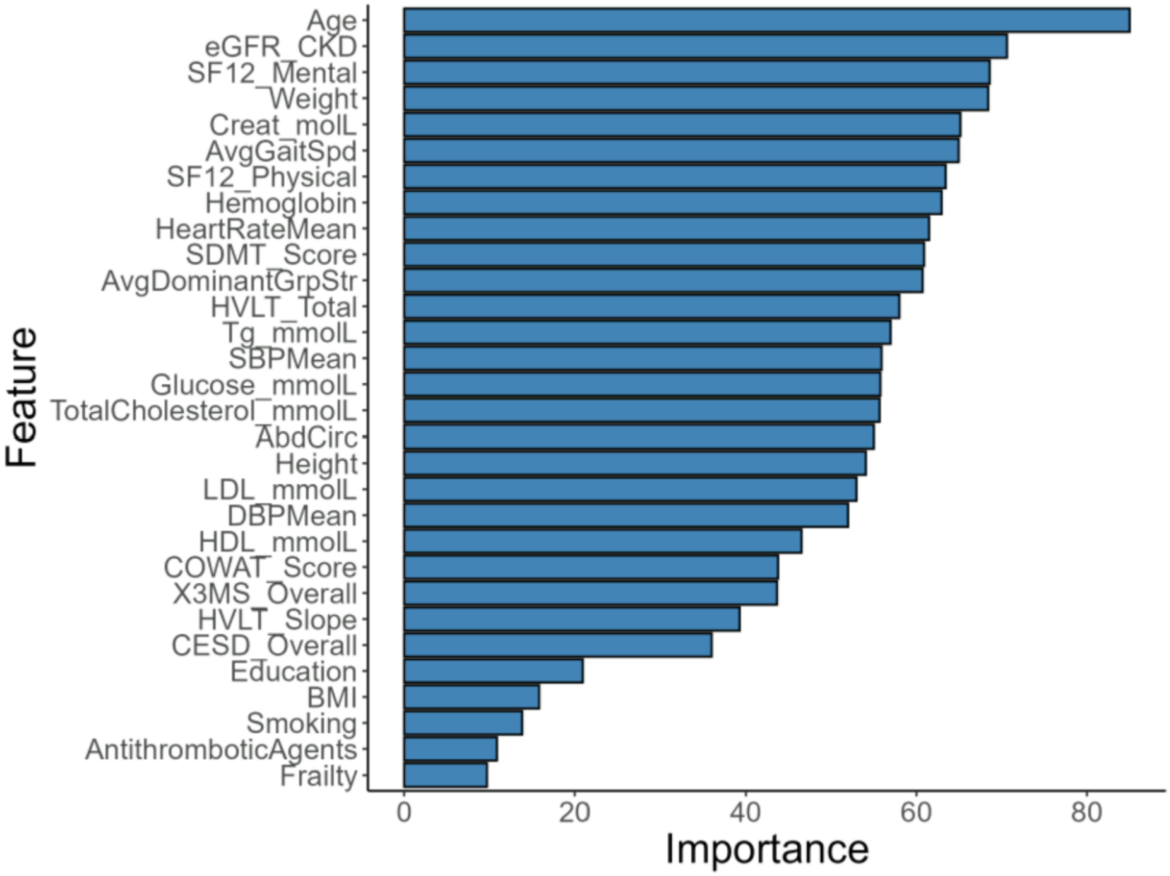


**Figure B.4:** Distribution of predicted bleeding risk scores based on the RSF model. Panel (a) shows the full risk distribution across all participants. Panel (b) shows the risk distribution stratified by treatment assignment.

a) Univariate

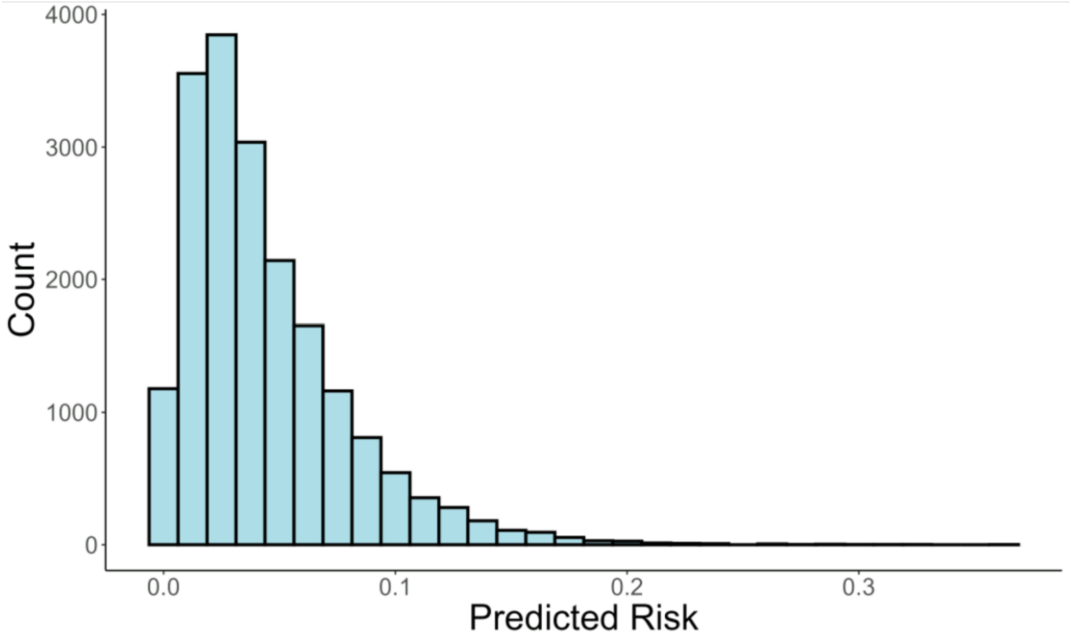

b) Stratified by treatment arm

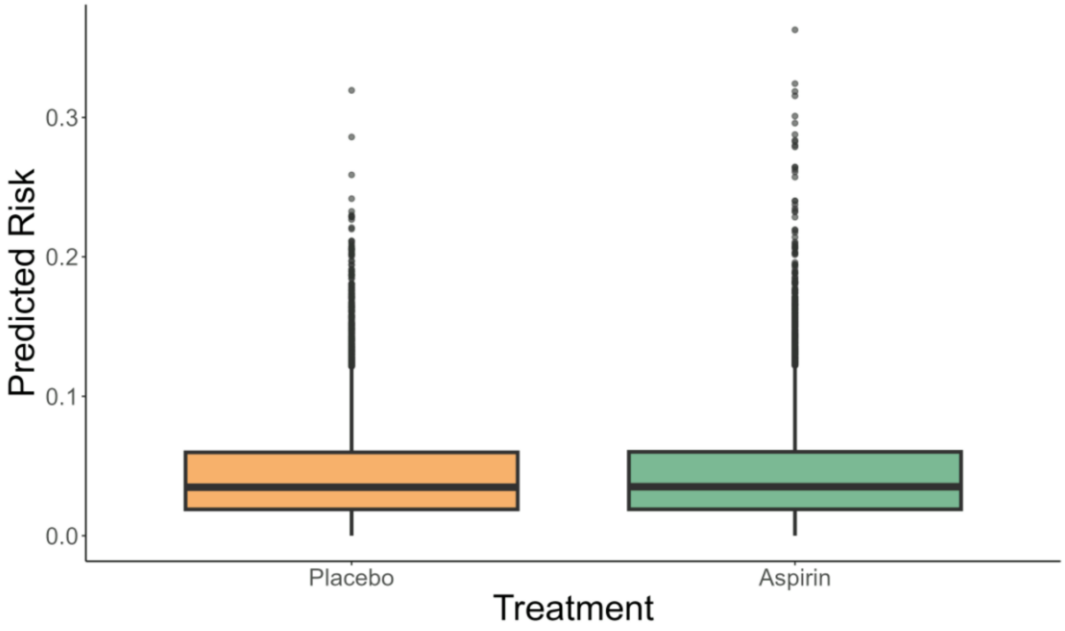


**Table B.2:** Summary statistics for predicted 5-year bleeding risk scores from the random survival forest model, stratified by RSF-based risk groups.

| Risk Group | N | Mean | SD | Min | Q1 | Median | Q3 | Max |
|---|---|---|---|---|---|---|---|---|
| Low | 3,823 | 0.009 | 0.005 | 0.000 | 0.005 | 0.009 | 0.013 | 0.016 |
| Intermediate | 11,468 | 0.037 | 0.014 | 0.016 | 0.025 | 0.035 | 0.048 | 0.068 |
| High | 3,823 | 0.101 | 0.033 | 0.068 | 0.077 | 0.091 | 0.114 | 0.363 |

**Table B.3:** Results from a Fine-Gray competing risks regression model evaluating the effect of aspirin versus placebo across RSF-derived bleeding risk groups. The model includes interaction terms to assess heterogeneity in treatment effect by risk group

| Term | Coef | exp(Coef) | SE | z | p-value | 2.5% CI | 97.5% CI |
|---|---|---|---|---|---|---|---|
| Treatment: Aspirin | 0.702 | 2.018 | 0.306 | 2.295 | 0.022 | 1.108 | 3.677 |
| RSF Risk Group: Intermediate | 0.995 | 2.703 | 0.265 | 3.757 | <0.001 | 1.609 | 4.542 |
| RSF Risk Group: High | 2.005 | 7.429 | 0.266 | 7.534 | <0.001 | 4.410 | 12.518 |
| Interaction: Aspirin × Intermediate | -0.241 | 0.786 | 0.326 | -0.739 | 0.460 | 0.415 | 1.489 |
| Interaction: Aspirin × High | -0.651 | 0.522 | 0.332 | -1.962 | 0.050 | 0.272 | 0.999 |

**Figure B.5:** Hazard ratios and 95% confidence intervals for aspirin versus placebo within each RSF-derived bleeding risk group, estimated using a Fine-Gray competing risks model. Below the plot, the table summarizes group sizes and the number of clinically significant bleeding events by treatment arm.

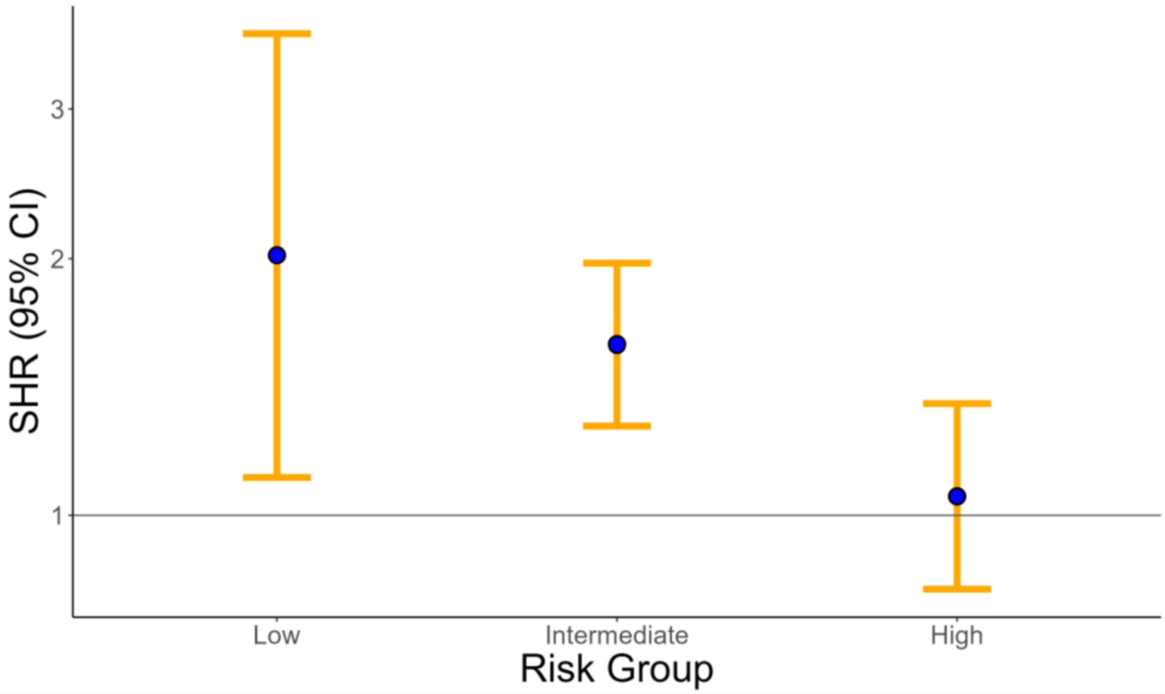


| Risk Group | n | n (Aspirin) | n (Placebo) | Bleeds (Aspirin) | Bleeds (Placebo) |
|---|---|---|---|---|---|
| Low | 3,823 | 1,901 | 1,922 | 32 | 16 |
| Intermediate | 11,468 | 5,709 | 5,759 | 204 | 131 |
| High | 3,823 | 1,915 | 1,908 | 125 | 118 |

**Table B.3:** Subdistribution hazard ratios and 95% confidence intervals for aspirin versus placebo within each RSF-based bleeding risk group, derived from a Fine-Gray competing risks model.

| RiskGroup | HR | HR CI Lower | HR CI Upper |
|---|---|---|---|
| Low | 2.018 | 1.108 | 3.677 |
| Intermediate | 1.586 | 1.274 | 1.976 |
| High | 1.053 | 0.818 | 1.354 |

**Table B.4:** Results from likelihood ratio and Wald tests evaluating the significance of treatment, RSF-derived bleeding risk groups, and their interaction in the Fine-Gray competing risks model.

| Test | Term | Risk Group | df | Statistic | p-value | Significance |
|---|---|---|---|---|---|---|
| Likelihood Ratio Test | Treatment:RiskGroupRSF2 | RiskGroupRSF2 | 2 | 7.5880 | 0.0225 | <0.05* |
| Wald Test | Treatment | RiskGroupRSF2 | 3 | 22.4192 | 5.336e-05 | <0.001*** |
| Wald Test | RiskGroupRSF2 | RiskGroupRSF2 | 2 | 96.3426 | 1.201e-21 | <0.001*** |
| Wald Test | Treatment:RiskGroupRSF2 | RiskGroupRSF2 | 2 | 7.5350 | 0.0231 | <0.05* |

**Table B.5:** Cumulative incidence of clinically significant bleeding at 5 years for aspirin and placebo groups, stratified by RSF-based baseline bleeding risk. Positive ARDs indicate increased bleeding risk with aspirin, and the number needed to harm (NNH) reflects the number of patients who would need to be treated with aspirin for one additional major bleeding event to occur.

| Risk Group | CIF (Aspirin) | CIF (Placebo) | ARD | Upper CI | Lower CI | NNH |
|---|---|---|---|---|---|---|
| Low | 0.0078 | 0.0054 | 0.0104 | 0.0023 | 0.0196 | 97 |
| Intermediate | 0.0221 | 0.0131 | 0.0142 | 0.0074 | 0.0208 | 71 |
| High | 0.0351 | 0.0348 | 0.0032 | -0.0142 | 0.0191 | 313 |

**Table B.6:** Results from a Q-test evaluating heterogeneity in ARDs between aspirin and placebo at 5 years across RSF-derived risk groups.

| Test Statistic | Degrees of Freedom | p-value | $I^2$ (%) | $H^2$ |
|---|---|---|---|---|
| Q = 1.5775 | 2 | 0.4544 | 0.00% | 0.79 |

**Figure B.6:** Cumulative incidence functions for clinically significant bleeding over time, stratified by RSF-based risk group.

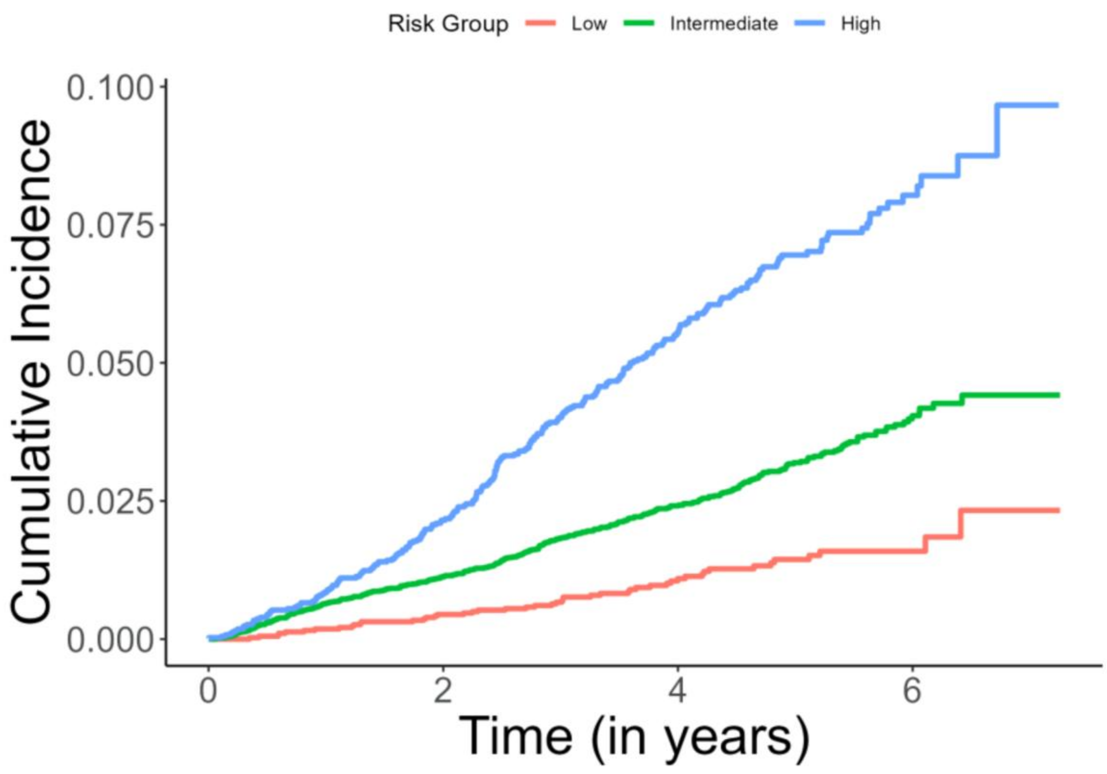

**Table B.7:** Cumulative incidence function values by group at evaluation times closest to 5 years, stratified by RSF-based risk group.

| Group | Eval_Time | CIF | CIF (%) |
|---|---|---|---|
| Low | 5.1170 | 0.0144 | 1.44% |
| Intermediate | 4.9856 | 0.0317 | 3.17% |
| High | 5.1006 | 0.0695 | 6.95% |

**Figure B.7:** Stratified cumulative incidence curves for clinically significant bleeding by treatment arm (aspirin vs. placebo) within each RSF-derived bleeding risk group.

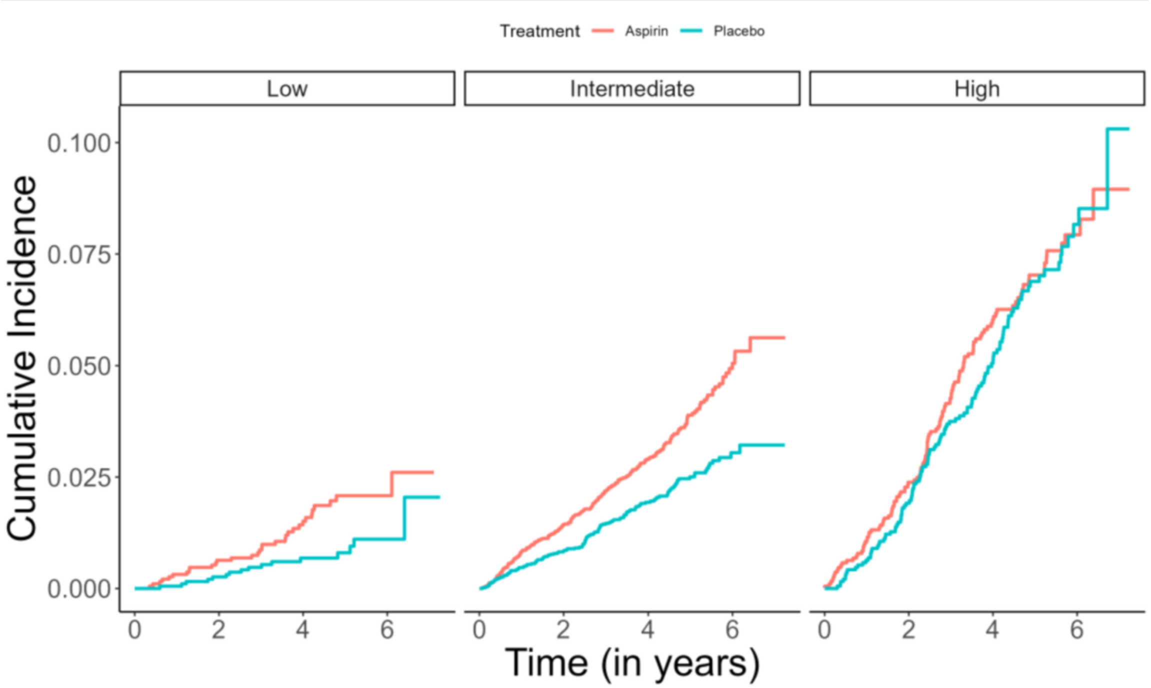

**Table B.8:** Cumulative incidence function estimates at approximately 5 years, stratified by treatment group, RSF-derived bleeding risk group, and event type.

| Treatment | Group | Cause | Eval_Time | CIF | CIF (%) |
|---|---|---|---|---|---|
| Placebo | Low | Main Outcome | 5.1170 | 0.0080 | 0.80% |
| Aspirin | Low | Main Outcome | 4.7912 | 0.0197 | 1.97% |
| Placebo | Intermediate | Main Outcome | 4.9856 | 0.0246 | 2.46% |
| Aspirin | Intermediate | Main Outcome | 5.0459 | 0.0389 | 3.89% |
| Placebo | High | Main Outcome | 5.1006 | 0.0688 | 6.88% |
| Aspirin | High | Main Outcome | 4.8597 | 0.0692 | 6.92% |
| Placebo | Low | Competing Risk | 3.9699 | 0.0114 | 1.14% |
| Aspirin | Low | Competing Risk | 5.0705 | 0.0118 | 1.18% |
| Placebo | Intermediate | Competing Risk | 4.9938 | 0.0404 | 4.04% |
| Aspirin | Intermediate | Competing Risk | 4.9938 | 0.0437 | 4.37% |

### Appendix C: Random Forest Additional Outputs

**Table C.1:** Summary of Random Forest model parameters used to estimate the probability of clinically significant bleeding.

| Parameter | Value |
|---|---|
| **Model type** | Probability estimation |
| **Number of trees** | 500 |
| **Sample size** | 19,114 |
| **Number of independent variables** | 116 |
| **mtry** (number of variables tried at each split) | 10 |
| **Target node size** | 10 |
| **Variable importance mode** | impurity |
| **Split rule** | gini |
| **OOB prediction error (Brier score)** | 0.0315 |

**Figure C.1:** Receiver operating characteristic (ROC) curve at 5 years for the random forest model predicting clinically significant bleeding.

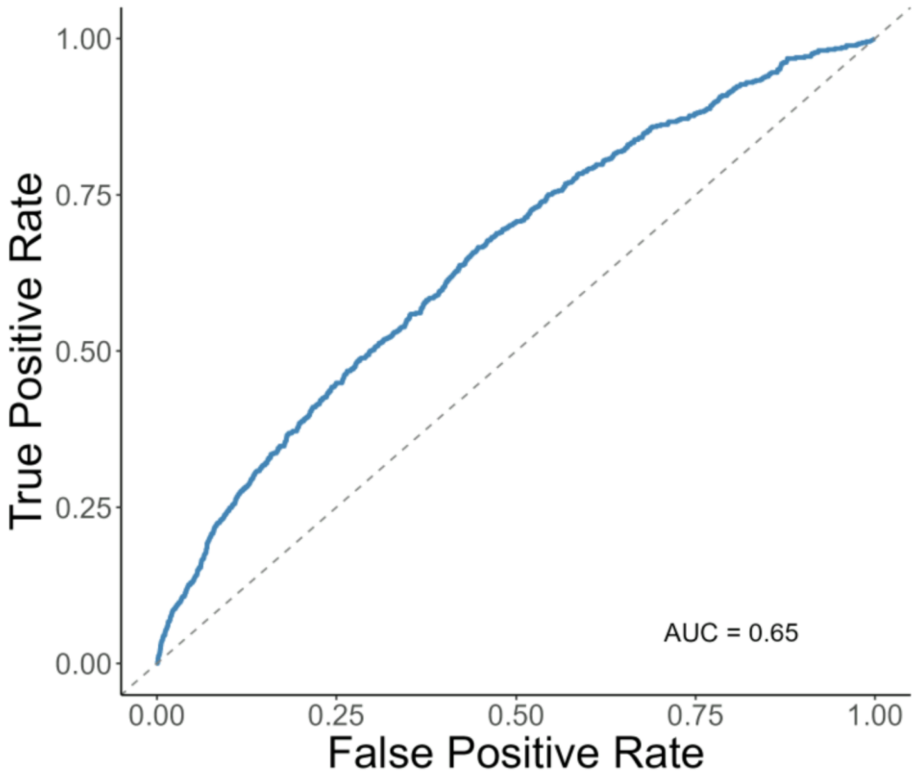

**Figure C.2:** Calibration plot for the Random Forest model predicting clinically significant bleeding risk. The plot compares observed event rates to predicted probabilities across deciles of predicted risk.

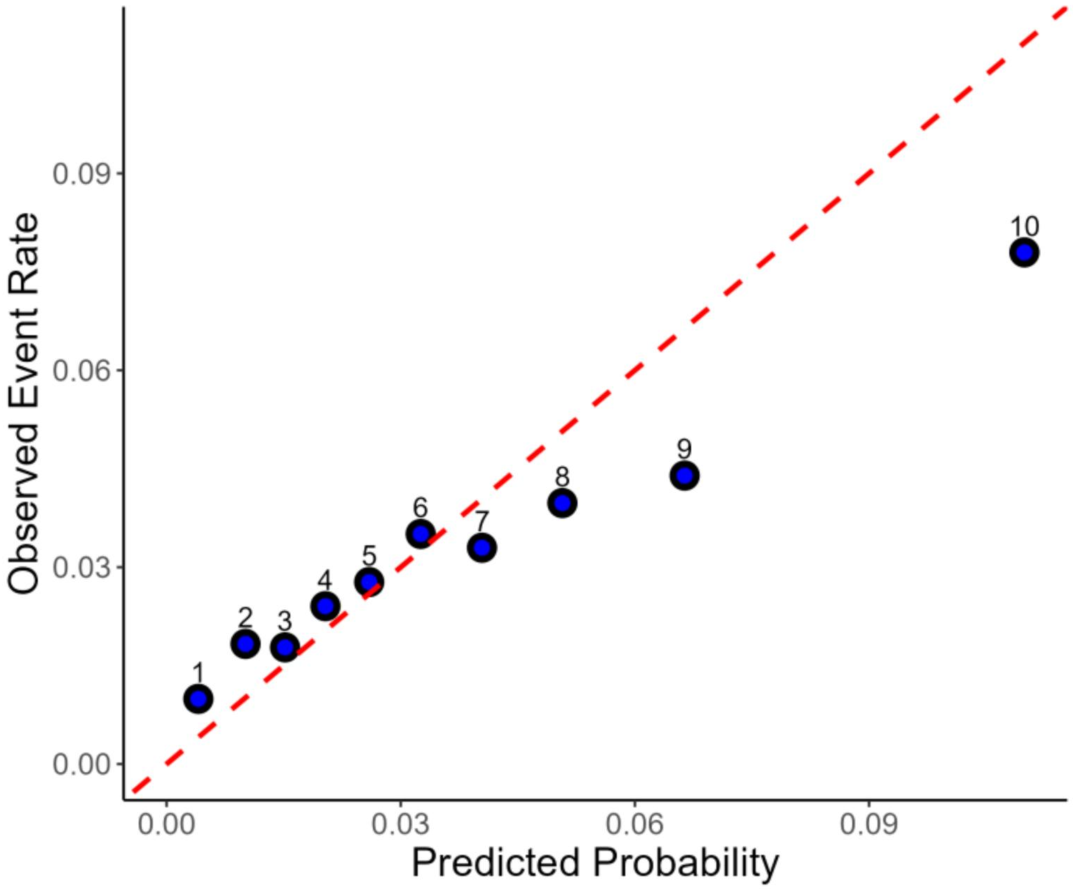

**Figure C.3:** Top predictors of clinically significant bleeding identified by the RF model using impurity-based variable importance. Higher values indicate greater contribution to the model's predictive accuracy.

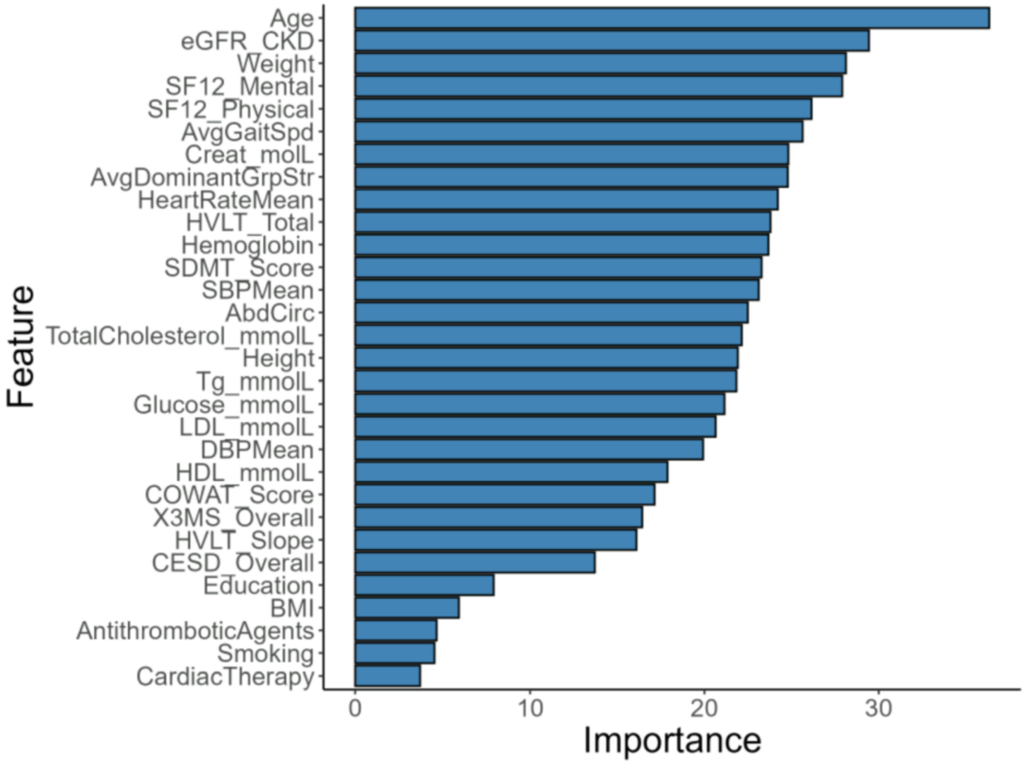

**Table C.2:** Results from a Fine-Gray competing risks regression model evaluating the effect of aspirin versus placebo across Random Forest–derived bleeding risk groups. The model includes interaction terms to assess heterogeneity in treatment effect by risk group.

| Term | Coef | exp(Coef) | SE | z | p-value | 2.5% CI | 97.5% CI |
|---|---|---|---|---|---|---|---|
| Treatment: Aspirin | 0.814 | 2.257 | 0.293 | 2.78 | 0.005 | 1.272 | 4.005 |
| RF Risk Group: Intermediate | 0.987 | 2.684 | 0.257 | 3.84 | <0.001 | 1.622 | 4.441 |
| RF Risk Group: High | 1.882 | 6.568 | 0.260 | 7.23 | <0.001 | 3.943 | 10.943 |
| Interaction: Aspirin × Intermediate | -0.408 | 0.665 | 0.313 | -1.30 | 0.191 | 0.360 | 1.228 |
| Interaction: Aspirin × High | -0.749 | 0.473 | 0.321 | -2.34 | 0.020 | 0.252 | 0.887 |

**Table C.3:** Results from likelihood ratio and Wald tests evaluating the significance of treatment, RF-derived bleeding risk groups, and their interaction in the Fine-Gray competing risks model.

| Test | Term | Risk Group | df | Statistic | p-value | Significance |
|---|---|---|---|---|---|---|
| Likelihood Ratio Test | Treatment:RiskGroupRF2 | RiskGroupRF2 | 2 | 7.3497 | 0.02535 | <0.05* |
| Wald Test | Treatment | RiskGroupRF2 | 3 | 21.4448 | 8.510e-05 | <0.001*** |
| Wald Test | RiskGroupRF2 | RiskGroupRF2 | 2 | 79.5940 | 5.204e-18 | <0.001*** |
| Wald Test | Treatment:RiskGroupRF2 | RiskGroupRF2 | 2 | 7.1995 | 0.02733 | <0.05* |

**Table C.4:** Results from a Q-test evaluating heterogeneity in absolute risk differences between aspirin and placebo at 5 years across Random Forest–derived bleeding risk groups.

| Test Statistic | Degrees of Freedom | p-value | $I^2$ (%) | $H^2$ |
|---|---|---|---|---|
| Q = 1.0183 | 2 | 0.6010 | 0.00% | 0.51 |

**Figure C.4:** Stratified cumulative incidence curves for clinically significant bleeding by treatment arm (aspirin vs. placebo) within each RF-derived bleeding risk group.

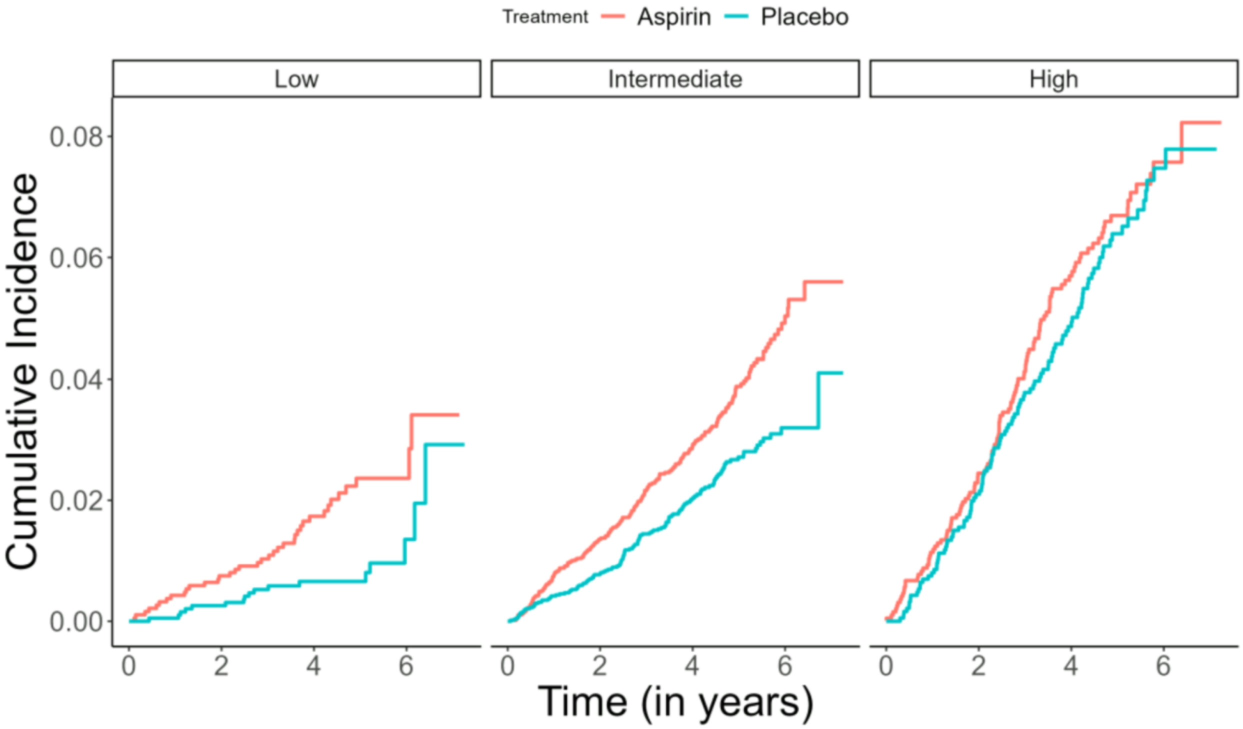

**Table C.5:** Cumulative incidence function estimates at approximately 5 years, stratified by treatment group, RF-derived bleeding risk group, and event type.

| Treatment | Group | Cause | Eval_Time | CIF | CIF (%) |
|---|---|---|---|---|---|
| Placebo | Low | Main Outcome | 5.1170 | 0.0066 | 0.66% |
| Aspirin | Low | Main Outcome | 4.9144 | 0.0223 | 2.23% |
| Placebo | Intermediate | Main Outcome | 4.9856 | 0.0267 | 2.67% |
| Aspirin | Intermediate | Main Outcome | 5.0459 | 0.0388 | 3.88% |
| Placebo | High | Main Outcome | 5.1006 | 0.0639 | 6.39% |
| Aspirin | High | Main Outcome | 4.8597 | 0.0659 | 6.59% |
| Placebo | Low | Competing Risk | 4.9665 | 0.0155 | 1.55% |
| Aspirin | Low | Competing Risk | 5.0705 | 0.0163 | 1.63% |
| Placebo | Intermediate | Competing Risk | 4.9637 | 0.0391 | 3.91% |
| Aspirin | Intermediate | Competing Risk | 5.0021 | 0.0439 | 4.39% |

### Appendix D: Predictor Variables and Descriptions

**Table D.1:** List of predictor variables and descriptions used in model development.

| Variable Name | Description |
|---|---|
| Age | Age in years |
| Gender | Self-reported gender |
| Country | Country of enrollment |
| Language | Language spoken |
| BornOverseas | Born outside native country |
| Education | Years of education completed |
| LivingSituation | Living situation (alone or with others) |
| RaceEthnicity | Self-identified race/ethnicity |
| Smoking | Current or former smoking status |
| AlcUse | Alcohol use |
| HistCancer | History of cancer |
| HistBowelPolyp | History of bowel polyps |
| HistDiabetes | History of diabetes |
| HistKidneyDisease | History of kidney disease |
| RegAspirin | Regular aspirin use prior to enrollment |

| Variable Name | Description |
|---|---|
| SBPMean | Mean systolic blood pressure |
| DBPMean | Mean diastolic blood pressure |
| HeartRateMean | Mean resting heart rate |
| IrregHB | Presence of irregular heartbeat |
| Height | Height (m) |
| Weight | Weight (kg) |
| AbdCirc | Abdominal circumference |
| BMI | Body mass index |
| Hemoglobin | Hemoglobin level (g/dL) |
| LDL_mmolL | LDL cholesterol (mmol/L) |
| HDL_mmolL | HDL cholesterol (mmol/L) |
| TG_mmolL | Triglycerides (mmol/L) |
| Creat_mmolL | Creatinine level (µmol/L) |
| Glucose_mmolL | Fasting blood glucose level (mmol/L) |
| TotalCholesterol_mmolL | Total cholesterol (mmol/L) |
| eGFR_CKD | Estimated glomerular filtration rate (chronic kidney disease formula) |

| Variable Name | Description |
|---|---|
| LipidLoweringAgents | Use of lipid-lowering medications |
| CalciumChannelBlockers | Use of calcium channel blockers |
| BetaBlockingAgents | Use of beta blockers |
| Diuretics | Use of diuretics |
| AntiInfl_AntiRheum_Prods | Use of anti-inflammatory or antirheumatic medications |
| AntiGoutPreps | Use of gout medications |
| Analgesics | Use of pain medications |
| ReninAngiotensinAgents | Use of ACE inhibitors or ARBs |
| AcidRelatedDrugs | Use of acid suppression therapy (e.g., PPIs) |
| BoneDiseaseDrugs | Use of osteoporosis-related medications |
| DermCorticosteroids | Use of topical corticosteroids |
| ConstipationDrugs | Use of constipation treatments |
| AntithromboticAgents | Use of antiplatelet or anticoagulant agents |
| Antiepileptics | Use of anti-seizure medications |
| Psychoanaleptics | Use of antidepressants or mood stabilizers |

| Variable Name | Description |
|---|---|
| Antibacterials | Use of antibiotics |
| CoughColdPreps | Use of cough and cold medications |
| Vitamins | Use of vitamin supplements |
| DiabetesDrugs | Use of diabetes medications |
| Ophthalmologicals | Use of eye medications |
| GIDisorderDrugs | Use of gastrointestinal disorder medications |
| Urologicals | Use of urologic medications |
| Antihistamines | Use of antihistamines |
| ThyroidTherapy | Use of thyroid hormone therapy |
| MuscleRelaxants | Use of muscle relaxant medications |
| AntineoplasticAgents | Use of cancer therapy drugs |
| Psycholeptics | Use of sedatives or antipsychotics |
| Corticosteroids | Use of systemic corticosteroids |
| OtherTherapeuticProds | Other therapeutic agents (unspecified) |
| Antihypertensives | Use of blood pressure lowering agents |

| Variable Name | Description |
|---|---|
| Antidiarrheals | Use of diarrhea treatment medications |
| SexHormones | Use of hormonal therapy |
| CardiacTherapy | Use of heart-related medications |
| OADDrugs | Use of oral anti-diabetic agents |
| Antifungals | Use of antifungal medications |
| EndocrineTherapy | Use of endocrine-modulating medications |
| NasalPreps | Use of nasal spray medications |
| EyeEarPreps | Use of ear or eye preparations |
| NervousSystemDrugs | Use of nervous system medications |
| AntiParkinsonDrugs | Use of medications for Parkinson's disease |
| Antiprotozoals | Use of anti-parasitic drugs |
| DermAntibiotics | Use of dermatological antibiotics |
| AntianemicPreps | Use of medications to treat anemia |
| ThroatPreps | Use of throat preparations (e.g., lozenges, sprays) |
| Otologicals | Use of ear-related medications |

| Variable Name | Description |
|---|---|
| Vasoprotectives | Use of vascular-protective agents |
| Antipsoriatics | Use of psoriasis treatments |
| MineralSupps | Use of mineral supplements |
| Immunosuppressants | Use of immunosuppressive therapy |
| Antivirals | Use of antiviral medications |
| Antipruritics | Use of anti-itch medications |
| DermPreps | Use of dermatological preparations |
| Antiemetics | Use of anti-nausea drugs |
| Gynecologicals | Use of gynecologic medications |
| CalciumHomeostasis | Disorders of calcium balance |
| FamHistHA | Family history of hypertension |
| FamHistStroke | Family history of stroke |
| FamHistDementia | Family history of dementia |
| FamHistKidneyDisease | Family history of kidney disease |
| FamHistKidneyDialysis | Family history of kidney dialysis |
| MotherHistHA | Mother's history of hypertension |

| Variable Name | Description |
|---|---|
| MotherHistStroke | Mother's history of stroke |
| MotherHistDementia | Mother's history of dementia |
| MotherHistKidneyDisease | Mother's history of kidney disease |
| MotherHistKidneyDialysis | Mother's history of kidney dialysis |
| FamHistCancer | Family history of cancer |
| MotherHistCancer | Mother's history of cancer |
| X3MS_Overall | Total score on 3MS cognitive screening test |
| COWAT_Score | Verbal fluency score (COWAT) |
| SDMT_Score | Symbol Digit Modalities Test score (cognitive speed) |
| HVLT_Total | Hopkins Verbal Learning Test total recall score |
| HVLT_Slope | Slope of learning across HVLT trials |
| AvgDominantGrpStr | Average dominant hand grip strength |
| AvgGaitSpd | Average gait speed (seconds/3 meters) |
| Any_ADL_Difficulty | Difficulty with activities of daily living (ADL) |

| Variable Name | Description |
| --- | --- |
| Any_IADL_Difficulty | Difficulty with instrumental activities of daily living (IADL) |
| Any_Mobility_Difficulty | Difficulty with performing mobility-related activities |
| CESD_Overall | Center for Epidemiologic Studies Depression Scale total score |
| Hypertension | Diagnosis of hypertension |
| Diabetes | Diagnosis of diabetes |
| Frailty | Frailty status based on clinical criteria |
| Dyslipidemia | Diagnosis of lipid disorders |
| SF12_Mental | Mental component summary score from SF-12 |
| SF12_Physical | Physical component summary score from SF-12 |
| Polypharmacy | Use of multiple concurrent medications |

### Appendix E: Pre-Imputation Missing Value Summary

**Table E.1:** Number of missing values for each feature prior to imputation. Many features with identical missingness (n = 868) were medication-related and are grouped under a single row labeled Medication variables.

| Feature | Missing |
|---|---|
| Medication variables* | 868 |
| LDL_mmolL | 502 |
| eGFR_CKD | 464 |
| Creat_molL | 464 |
| HDL_mmolL | 446 |
| Glucose_mmolL | 359 |
| AvgDominantGrpStr | 286 |
| Tg_mmolL | 218 |
| AbdCirc | 209 |
| TotalCholesterol_mmolL | 197 |
| HVLT_Total | 111 |
| HVLT_Slope | 107 |
| AvgGaitSpd | 96 |
| BMI | 89 |
| SDMT_Score | 84 |

| | |
|---|---|
| Weight | 61 |
| Height | 48 |
| COWAT_Score | 31 |
| HistDiabetes | 30 |
| HistKidneyDisease | 29 |
| HistCancer | 22 |
| HistBowelPolyp | 16 |
| SF12_Mental | 8 |
| SF12_Physical | 8 |
| FamHistCancer | 6 |
| HeartRateMean | 5 |
| MotherHistCancer | 4 |
| CESD_Overall | 4 |
| IrregHB | 3 |
| RegAspirin | 2 |
| Hemoglobin | 2 |
| FamHistHA | 2 |
| FamHistStroke | 2 |
| FamHistDementia | 2 |

| | |
|---|---|
| FamHistKidneyDisease | 2 |
| FamHistKidneyDialysis | 2 |
| MotherHistHA | 2 |
| MotherHistStroke | 2 |
| MotherHistDementia | 2 |
| MotherHistKidneyDisease | 2 |
| MotherHistKidneyDialysis | 2 |
| Education | 1 |

### Appendix F: Baseline Characteristics of ASPREE Participants by Treatment Group

**Table F.1:** Summary of demographic, clinical, lifestyle, and medication-related baseline characteristics of ASPREE participants, stratified by treatment assignment.

| | Stratified by Treatment | | |
|---|---|---|---|
| | **Placebo** | **Aspirin** | **p** |
| n | 9,589 | 9,525 | |
| Age (mean (SD)) | 75.07 (4.51) | 75.16 (4.58) | 0.171 |
| Gender = Female (%) | 5,409 (56.4) | 5,373 (56.4) | 1.000 |
| Country = US (%) | 1,208 (12.6) | 1,203 (12.6) | 0.964 |
| Language = Non-english (%) | 401 ( 4.2) | 450 ( 4.7) | 0.075 |
| BornOverseas = Yes (%) | 2,133 (22.2) | 2,143 (22.5) | 0.686 |
| Education (%) | | | 0.550 |
| <9 yrs | 1,467 (15.3) | 1,535 (16.1) | |
| 9-11 yrs | 2,862 (29.8) | 2,772 (29.1) | |
| 12 yrs | 1,154 (12.0) | 1,165 (12.2) | |
| 13-15 yrs | 1,618 (16.9) | 1,637 (17.2) | |
| 16 yrs | 894 ( 9.3) | 872 ( 9.2) | |
| 17-21 yrs | 1,594 (16.6) | 1,544 (16.2) | |
| LivingSituation (%) | | | 0.289 |
| At home alone | 3,154 (32.9) | 3,097 (32.5) | |
| At home with family, friends or spouse | 6,392 (66.7) | 6,388 (67.1) | |
| In a residential home (supervised care) | 43 ( 0.4) | 37 ( 0.4) | |
| In a nursing home alone | 0 ( 0.0) | 3 ( 0.0) | |
| RaceEthnicity (%) | | | 0.559 |
| Black | 450 ( 4.7) | 451 ( 4.7) | |
| Hispanic | 248 ( 2.6) | 240 ( 2.5) | |
| Other | 150 ( 1.6) | 126 ( 1.3) | |
| White | 8,741 (91.2) | 8,708 (91.4) | |
| Smoking (%) | | | 0.498 |
| Current | 383 ( 4.0) | 352 ( 3.7) | |
| Former | 3,890 (40.6) | 3,909 (41.0) | |
| Never | 5,316 (55.4) | 5,264 (55.3) | |
| AlcUse (%) | | | 0.892 |
| Current | 7,333 (76.5) | 7,309 (76.7) | |
| Former | 570 ( 5.9) | 566 ( 5.9) | |
| Never | 1,686 (17.6) | 1,650 (17.3) | |
| HistCancer = Yes (%) | 1,834 (19.1) | 1,831 (19.2) | 0.879 |
| HistBowelPolyp = Yes (%) | 2,000 (20.9) | 1,896 (19.9) | 0.106 |
| HistDiabetes = Yes (%) | 844 ( 8.8) | 865 ( 9.1) | 0.514 |
| HistKidneyDisease = Yes (%) | 201 ( 2.1) | 238 ( 2.5) | 0.070 |
| RegAspirin = Yes (%) | 1,042 (10.9) | 1,053 (11.1) | 0.694 |

| | | | |
|---|---|---|---|
| SBPMean (mean (SD)) | 139.22 (16.49) | 139.16 (16.53) | 0.806 |
| DBPMean (mean (SD)) | 77.19 (9.93) | 77.34 (10.03) | 0.294 |
| HeartRateMean (mean (SD)) | 70.87 (10.82) | 70.58 (10.63) | 0.065 |
| IrregHB = Yes (%) | 203 ( 2.1) | 189 ( 2.0) | 0.551 |
| Height (mean (SD)) | 1.65 (0.09) | 1.65 (0.09) | 0.323 |
| Weight (mean (SD)) | 77.02 (14.78) | 76.94 (15.14) | 0.732 |
| AbdCirc (mean (SD)) | 97.20 (12.75) | 97.10 (12.97) | 0.593 |
| BMI (%) | | | 0.708 |
| Underweight | 48 ( 0.5) | 55 ( 0.6) | |
| Normal | 2,422 (25.3) | 2,458 (25.8) | |
| Overweight | 4,295 (44.8) | 4,217 (44.3) | |
| Obese | 2,824 (29.5) | 2,795 (29.3) | |
| Hemoglobin (mean (SD)) | 14.17 (1.22) | 14.15 (1.22) | 0.190 |
| LDL_mmolL (mean (SD)) | 3.05 (0.87) | 3.04 (0.88) | 0.739 |
| HDL_mmolL (mean (SD)) | 1.58 (0.46) | 1.58 (0.46) | 0.817 |
| Tg_mmolL (mean (SD)) | 1.33 (0.65) | 1.33 (0.67) | 0.978 |
| Creat_molL (mean (SD)) | 80.03 (19.02) | 80.10 (19.98) | 0.791 |
| Glucose_mmolL (mean (SD)) | 5.49 (1.06) | 5.48 (1.04) | 0.852 |
| TotalCholesterol_mmolL (mean (SD)) | 5.24 (0.98) | 5.23 (0.99) | 0.617 |
| eGFR_CKD (mean (SD)) | 72.88 (13.89) | 72.89 (13.98) | 0.950 |
| LipidLoweringAgents (%) | | | 0.791 |
| No | 4,940 (51.5) | 4,951 (52.0) | |
| Yes | 4,208 (43.9) | 4,147 (43.5) | |
| Not recorded | 441 ( 4.6) | 427 ( 4.5) | |
| CalciumChannelBlockers (%) | | | 0.009 |
| No | 6,747 (70.4) | 6,527 (68.5) | |
| Yes | 2,401 (25.0) | 2,571 (27.0) | |
| Not recorded | 441 ( 4.6) | 427 ( 4.5) | |
| BetaBlockingAgents (%) | | | 0.555 |
| No | 7,616 (79.4) | 7,523 (79.0) | |
| Yes | 1,532 (16.0) | 1,575 (16.5) | |
| Not recorded | 441 ( 4.6) | 427 ( 4.5) | |
| Diuretics (%) | | | 0.238 |
| No | 6,648 (69.3) | 6,512 (68.4) | |
| Yes | 2,500 (26.1) | 2,586 (27.1) | |
| Not recorded | 441 ( 4.6) | 427 ( 4.5) | |
| AntiInfl_AntiRheum_Prods (%) | | | 0.924 |
| No | 6,636 (69.2) | 6,594 (69.2) | |
| Yes | 2,512 (26.2) | 2,504 (26.3) | |
| Not recorded | 441 ( 4.6) | 427 ( 4.5) | |
| AntiGoutPreps (%) | | | 0.421 |
| No | 8,630 (90.0) | 8,543 (89.7) | |
| Yes | 518 ( 5.4) | 555 ( 5.8) | |
| Not recorded | 441 ( 4.6) | 427 ( 4.5) | |
| Analgestics (%) | | | 0.926 |

| | | | |
|---|---|---|---|
| No | 5,510 (57.5) | 5,485 (57.6) | |
| Yes | 3,638 (37.9) | 3,613 (37.9) | |
| Not recorded | 441 ( 4.6) | 427 ( 4.5) | |
| ReninAngiotensinAgents (%) | | | 0.582 |
| No | 4,086 (42.6) | 3,999 (42.0) | |
| Yes | 5,062 (52.8) | 5,099 (53.5) | |
| Not recorded | 441 ( 4.6) | 427 ( 4.5) | |
| AcidRelatedDrugs (%) | | | 0.658 |
| No | 5,539 (57.8) | 5,454 (57.3) | |
| Yes | 3,609 (37.6) | 3,644 (38.3) | |
| Not recorded | 441 ( 4.6) | 427 ( 4.5) | |
| BoneDiseaseDrugs (%) | | | 0.915 |
| No | 7,998 (83.4) | 7,962 (83.6) | |
| Yes | 1,150 (12.0) | 1,136 (11.9) | |
| Not recorded | 441 ( 4.6) | 427 ( 4.5) | |
| DermCorticosteroids (%) | | | 0.624 |
| No | 8,222 (85.7) | 8,213 (86.2) | |
| Yes | 926 ( 9.7) | 885 ( 9.3) | |
| Not recorded | 441 ( 4.6) | 427 ( 4.5) | |
| ConstipationDrugs (%) | | | 0.137 |
| No | 8,973 (93.6) | 8,886 (93.3) | |
| Yes | 175 ( 1.8) | 212 ( 2.2) | |
| Not recorded | 441 ( 4.6) | 427 ( 4.5) | |
| AntithromboticAgents (%) | | | 0.009 |
| No | 7,311 (76.2) | 7,433 (78.0) | |
| Yes | 1,837 (19.2) | 1,665 (17.5) | |
| Not recorded | 441 ( 4.6) | 427 ( 4.5) | |
| Antiepileptics (%) | | | 0.392 |
| No | 8,362 (87.2) | 8,266 (86.8) | |
| Yes | 786 ( 8.2) | 832 ( 8.7) | |
| Not recorded | 441 ( 4.6) | 427 ( 4.5) | |
| Psychoanaleptics (%) | | | 0.872 |
| No | 7,322 (76.4) | 7,301 (76.7) | |
| Yes | 1,826 (19.0) | 1,797 (18.9) | |
| Not recorded | 441 ( 4.6) | 427 ( 4.5) | |
| Antibacterials (%) | | | 0.664 |
| No | 8,225 (85.8) | 8,213 (86.2) | |
| Yes | 923 ( 9.6) | 885 ( 9.3) | |
| Not recorded | 441 ( 4.6) | 427 ( 4.5) | |
| CoughColdPreps (%) | | | 0.017 |
| No | 8,630 (90.0) | 8,491 (89.1) | |
| Yes | 518 ( 5.4) | 607 ( 6.4) | |
| Not recorded | 441 ( 4.6) | 427 ( 4.5) | |
| Vitamins (%) | | | 0.910 |
| No | 7,352 (76.7) | 7,301 (76.7) | |

| | | | |
|---|---|---|---|
| Yes | 1,796 (18.7) | 1,797 (18.9) | |
| Not recorded | 441 ( 4.6) | 427 ( 4.5) | |
| DiabetesDrugs (%) | | | 0.389 |
| No | 8,373 (87.3) | 8,277 (86.9) | |
| Yes | 775 ( 8.1) | 821 ( 8.6) | |
| Not recorded | 441 ( 4.6) | 427 ( 4.5) | |
| Ophthalmologicals (%) | | | 0.169 |
| No | 8,233 (85.9) | 8,112 (85.2) | |
| Yes | 915 ( 9.5) | 986 (10.4) | |
| Not recorded | 441 ( 4.6) | 427 ( 4.5) | |
| GIDisorderDrugs (%) | | | 0.673 |
| No | 8,864 (92.4) | 8,834 (92.7) | |
| Yes | 284 ( 3.0) | 264 ( 2.8) | |
| Not recorded | 441 ( 4.6) | 427 ( 4.5) | |
| Urologicals (%) | | | 0.900 |
| No | 8,152 (85.0) | 8,097 (85.0) | |
| Yes | 996 (10.4) | 1,001 (10.5) | |
| Not recorded | 441 ( 4.6) | 427 ( 4.5) | |
| Antihistamines (%) | | | 0.275 |
| No | 8,783 (91.6) | 8,775 (92.1) | |
| Yes | 365 ( 3.8) | 323 ( 3.4) | |
| Not recorded | 441 ( 4.6) | 427 ( 4.5) | |
| ThyroidTherapy (%) | | | 0.705 |
| No | 8,254 (86.1) | 8,179 (85.9) | |
| Yes | 894 ( 9.3) | 919 ( 9.6) | |
| Not recorded | 441 ( 4.6) | 427 ( 4.5) | |
| MuscleRelaxants (%) | | | 0.842 |
| No | 9,075 (94.6) | 9,020 (94.7) | |
| Yes | 73 ( 0.8) | 78 ( 0.8) | |
| Not recorded | 441 ( 4.6) | 427 ( 4.5) | |
| AntineoplasticAgents (%) | | | 0.883 |
| No | 8,852 (92.3) | 8,796 (92.3) | |
| Yes | 296 ( 3.1) | 302 ( 3.2) | |
| Not recorded | 441 ( 4.6) | 427 ( 4.5) | |
| Psycholeptics (%) | | | 0.918 |
| No | 7,606 (79.3) | 7,572 (79.5) | |
| Yes | 1,542 (16.1) | 1,526 (16.0) | |
| Not recorded | 441 ( 4.6) | 427 ( 4.5) | |
| Cortisosteroids (%) | | | 0.901 |
| No | 8,366 (87.2) | 8,311 (87.3) | |
| Yes | 782 ( 8.2) | 787 ( 8.3) | |
| Not recorded | 441 ( 4.6) | 427 ( 4.5) | |
| OtherTherapeuticProds (%) | | | 0.496 |
| No | 8997 (93.8) | 8928 (93.7) | |
| Yes | 151 ( 1.6) | 170 ( 1.8) | |

| | | | |
|---|---|---|---|
| Not recorded | 441 ( 4.6) | 427 ( 4.5) | |
| Antihypertensives (%) | | | 0.922 |
| No | 8,674 (90.5) | 8,623 (90.5) | |
| Yes | 474 ( 4.9) | 475 ( 5.0) | |
| Not recorded | 441 ( 4.6) | 427 ( 4.5) | |
| Antidiarrheals (%) | | | 0.229 |
| No | 8,933 (93.2) | 8,917 (93.6) | |
| Yes | 215 ( 2.2) | 181 ( 1.9) | |
| Not recorded | 441 ( 4.6) | 427 ( 4.5) | |
| SexHormones (%) | | | 0.124 |
| No | 8,390 (87.5) | 8,417 (88.4) | |
| Yes | 758 ( 7.9) | 681 ( 7.1) | |
| Not recorded | 441 ( 4.6) | 427 ( 4.5) | |
| CardiacTherapy (%) | | | 0.918 |
| No | 8,778 (91.5) | 8,734 (91.7) | |
| Yes | 370 ( 3.9) | 364 ( 3.8) | |
| Not recorded | 441 ( 4.6) | 427 ( 4.5) | |
| OADDrugs (%) | | | 0.544 |
| No | 7,340 (76.5) | 7,355 (77.2) | |
| Yes | 1,808 (18.9) | 1,743 (18.3) | |
| Not recorded | 441 ( 4.6) | 427 ( 4.5) | |
| Antifungals (%) | | | 0.295 |
| No | 9,013 (94.0) | 8,938 (93.8) | |
| Yes | 135 ( 1.4) | 160 ( 1.7) | |
| Not recorded | 441 ( 4.6) | 427 ( 4.5) | |
| EndocrineTherapy (%) | | | 0.668 |
| No | 8,923 (93.1) | 8,857 (93.0) | |
| Yes | 225 ( 2.3) | 241 ( 2.5) | |
| Not recorded | 441 ( 4.6) | 427 ( 4.5) | |
| NasalPreps (%) | | | 0.923 |
| No | 9,031 (94.2) | 8,980 (94.3) | |
| Yes | 117 ( 1.2) | 118 ( 1.2) | |
| Not recorded | 441 ( 4.6) | 427 ( 4.5) | |
| EyeEarPreps (%) | | | 0.803 |
| No | 9,080 (94.7) | 9,024 (94.7) | |
| Yes | 68 ( 0.7) | 74 ( 0.8) | |
| Not recorded | 441 ( 4.6) | 427 ( 4.5) | |
| NervousSystemDrugs (%) | | | 0.888 |
| No | 9,036 (94.2) | 8,991 (94.4) | |
| Yes | 112 ( 1.2) | 107 ( 1.1) | |
| Not recorded | 441 ( 4.6) | 427 ( 4.5) | |
| AntiParkinsonDrugs (%) | | | 0.205 |
| No | 8,912 (92.9) | 8,899 (93.4) | |
| Yes | 236 ( 2.5) | 199 ( 2.1) | |
| Not recorded | 441 ( 4.6) | 427 ( 4.5) | |

| | | | |
|---|---|---|---|
| Antiprotozoals (%) | | | 0.341 |
| No | 8,975 (93.6) | 8,899 (93.4) | |
| Yes | 173 ( 1.8) | 199 ( 2.1) | |
| Not recorded | 441 ( 4.6) | 427 ( 4.5) | |
| DermAntibiotics (%) | | | 0.328 |
| No | 9,015 (94.0) | 8,988 (94.4) | |
| Yes | 133 ( 1.4) | 110 ( 1.2) | |
| Not recorded | 441 ( 4.6) | 427 ( 4.5) | |
| AntianemicPreps (%) | | | 0.157 |
| No | 8,821 (92.0) | 8,724 (91.6) | |
| Yes | 327 ( 3.4) | 374 ( 3.9) | |
| Not recorded | 441 ( 4.6) | 427 ( 4.5) | |
| ThroatPreps (%) | | | 0.923 |
| No | 9,063 (94.5) | 9,012 (94.6) | |
| Yes | 85 ( 0.9) | 86 ( 0.9) | |
| Not recorded | 441 ( 4.6) | 427 ( 4.5) | |
| Otologicals (%) | | | 0.866 |
| No | 9,069 (94.6) | 9,024 (94.7) | |
| Yes | 79 ( 0.8) | 74 ( 0.8) | |
| Not recorded | 441 ( 4.6) | 427 ( 4.5) | |
| Vasoprotectives (%) | | | 0.474 |
| No | 9,141 (95.3) | 9,086 (95.4) | |
| Yes | 7 ( 0.1) | 12 ( 0.1) | |
| Not recorded | 441 ( 4.6) | 427 ( 4.5) | |
| Antipsoriatics (%) | | | 0.809 |
| No | 9,086 (94.8) | 9,042 (94.9) | |
| Yes | 62 ( 0.6) | 56 ( 0.6) | |
| Not recorded | 441 ( 4.6) | 427 ( 4.5) | |
| MineralSupps (%) | | | 0.815 |
| No | 8,914 (93.0) | 8,876 (93.2) | |
| Yes | 234 ( 2.4) | 222 ( 2.3) | |
| Not recorded | 441 ( 4.6) | 427 ( 4.5) | |
| Immunosuppressants (%) | | | 0.557 |
| No | 9,078 (94.7) | 9,016 (94.7) | |
| Yes | 70 ( 0.7) | 82 ( 0.9) | |
| Not recorded | 441 ( 4.6) | 427 ( 4.5) | |
| Antivirals (%) | | | 0.456 |
| No | 9,065 (94.5) | 9,030 (94.8) | |
| Yes | 83 ( 0.9) | 68 ( 0.7) | |
| Not recorded | 441 ( 4.6) | 427 ( 4.5) | |
| Antipruritics (%) | | | 0.823 |
| No | 9,129 (95.2) | 9,082 (95.3) | |
| Yes | 19 ( 0.2) | 16 ( 0.2) | |
| Not recorded | 441 ( 4.6) | 427 ( 4.5) | |
| DermPreps (%) | | | 0.914 |

| | | | |
|---|---|---|---|
| No | 9,129 (95.2) | 9,078 (95.3) | |
| Yes | 19 ( 0.2) | 20 ( 0.2) | |
| Not recorded | 441 ( 4.6) | 427 ( 4.5) | |
| Antiemetics (%) | | | 0.924 |
| No | 9,115 (95.1) | 9,066 (95.2) | |
| Yes | 33 ( 0.3) | 32 ( 0.3) | |
| Not recorded | 441 ( 4.6) | 427 ( 4.5) | |
| Gynecologicals (%) | | | 0.784 |
| No | 9,147 (95.4) | 9,096 (95.5) | |
| Yes | 1 ( 0.0) | 2 ( 0.0) | |
| Not recorded | 441 ( 4.6) | 427 ( 4.5) | |
| CalciumHomeostasis (%) | | | 0.909 |
| No | 9,137 (95.3) | 9,088 (95.4) | |
| Yes | 11 ( 0.1) | 10 ( 0.1) | |
| Not recorded | 441 ( 4.6) | 427 ( 4.5) | |
| FamHistHA = Yes (%) | 2,414 (25.2) | 2,328 (24.4) | 0.247 |
| FamHistStroke = Yes (%) | 1,212 (12.6) | 1,197 (12.6) | 0.897 |
| FamHistDementia = Yes (%) | 561 ( 5.9) | 591 ( 6.2) | 0.318 |
| FamHistKidneyDisease = Yes (%) | 171 ( 1.8) | 176 ( 1.8) | 0.780 |
| FamHistKidneyDialysis = Yes (%) | 19 ( 0.2) | 14 ( 0.1) | 0.498 |
| MotherHistHA = Yes (%) | 1,443 (15.0) | 1,434 (15.1) | 1.000 |
| MotherHistStroke = Yes (%) | 1,576 (16.4) | 1,564 (16.4) | 0.992 |
| MotherHistDementia = Yes (%) | 1,610 (16.8) | 1,606 (16.9) | 0.911 |
| MotherHistKidneyDisease = Yes (%) | 235 ( 2.5) | 241 ( 2.5) | 0.760 |
| MotherHistKidneyDialysis = Yes (%) | 21 ( 0.2) | 35 ( 0.4) | 0.078 |
| FamHistCancer = Yes (%) | 2,216 (23.1) | 2,172 (22.8) | 0.626 |
| MotherHistCancer = Yes (%) | 2,113 (22.0) | 2,072 (21.8) | 0.649 |
| X3MS_Overall (mean (SD)) | 93.46 (4.59) | 93.39 (4.65) | 0.264 |
| COWAT_Score (mean (SD)) | 12.11 (4.55) | 12.08 (4.58) | 0.621 |
| SDMT_Score (mean (SD)) | 36.80 (10.20) | 36.67 (10.09) | 0.375 |
| HVLT_Total (mean (SD)) | 30.21 (7.92) | 30.20 (7.99) | 0.908 |
| HVLT_Slope (mean (SD)) | 2.15 (2.23) | 2.13 (2.26) | 0.515 |
| AvgDominantGrpStr (mean (SD)) | 26.96 (10.02) | 26.80 (10.01) | 0.268 |
| AvgGaitSpd (mean (SD)) | 3.16 (0.96) | 3.15 (0.94) | 0.577 |
| Any_ADL_Difficulty = Yes (%) | 915 ( 9.5) | 909 ( 9.5) | 1.000 |
| Any_IADL_Difficulty = Yes (%) | 1,135 (11.8) | 1,148 (12.1) | 0.661 |
| Any_Mobility_Difficulty = Yes (%) | 4,702 (49.0) | 4,710 (49.4) | 0.577 |
| CESD_Overall (mean (SD)) | 3.22 (3.30) | 3.15 (3.30) | 0.133 |
| MajorHem = Yes (%) | 265 ( 2.8) | 361 ( 3.8) | <0.001 |
| MajorHem_YSR (mean (SD)) | 4.43 (1.39) | 4.41 (1.40) | 0.251 |
| CompPrimEnd (mean (SD)) | 0.10 (0.29) | 0.10 (0.30) | 0.747 |
| CompPrimEnd_YSR (mean (SD)) | 4.50 (1.34) | 4.50 (1.34) | 0.749 |
| Hypertension = Yes (%) | 7,140 (74.5) | 7,055 (74.1) | 0.546 |
| Diabetes = Yes (%) | 1,021 (10.6) | 1,024 (10.8) | 0.836 |
| Frailty (%) | | | 0.875 |

| | | | |
|---|---|---|---|
| Not frail | 5,643 (58.8) | 5,603 (58.8) | |
| Pre-frail | 3,740 (39.0) | 3,707 (38.9) | |
| Frail | 206 ( 2.1) | 215 ( 2.3) | |
| Dyslipidemia (mean (SD)) | 0.66 (0.47) | 0.65 (0.48) | 0.117 |
| SF12_Mental (mean (SD)) | 55.60 (7.16) | 55.75 (7.10) | 0.136 |
| SF12_Physical (mean (SD)) | 48.38 (8.78) | 48.29 (8.74) | 0.482 |
| Polypharmacy = Yes (%) | 2,484 (25.9) | 2,604 (27.3) | 0.026 |
